# Exploration of the circulating human secretome through protein quantitative trait analysis identifies an association between circulating levels of apolipoprotein L1 and risk of giant cell arteritis

**DOI:** 10.1101/2024.01.19.24301534

**Authors:** NJM Chaddock, M Zulcinski, J Martin, A Mälarstig, JE Peters, MM Iles, AW Morgan UK GCA Consortium

**Author notes:** **Corresponding author:** Ann W Morgan, Leeds Institute of Cardiovascular and Metabolic Medicine, School of Medicine, LIGHT Building, Clarendon Way, University of Leeds, Leeds LS2 9DA, UK. See Appendix 1 for members of the UK GCA Consortium. **Apolipoprotein-L1 association with giant cell arteritis**.

## Abstract

**Background:** Glucocorticoid monotherapy remains the principal treatment for giant cell arteritis (GCA), yet concurrent toxicity and adverse effects highlight the need for targeted therapies and improved risk stratification. Previous work suggests that evidence of genetic association can improve success rates in clinical trials and identify biomarkers for risk assessment, particularly when combined with other ‘omics data, such as proteomics. However, relatively little is currently known about the genetic basis of GCA.

**Methods:** Polygenic risk scores (PRS) were developed for 169 human plasma proteins and tested for association with GCA susceptibility (cases *N*=729, controls *N*=2,619). Associated PRS were replicated in an independent cohort (cases *N*=1,129, controls *N*=2,654) and their respective proteins were evaluated for causality using Mendelian randomization (MR). Finally, relationships between proteins with GCA-associated PRS were assessed using protein-protein interaction (PPI) network analysis

**Results:** The Apolipoprotein L1 (APOL1) PRS had a statistically significant GCA association with a protective effect (*P-value*[*P*]=1 x 10^-4^), which replicated in an independent dataset (*P*=8.69 x 10^-4^), and MR analysis supported a causal relationship (*beta*=-0.093; *SE*=0.02*; P*=4.42 x 10^-9^). PPI network analysis of proteins with GCA-associated PRS revealed enrichment for “negative regulation of fibrinolysis” and “negative regulation of blood coagulation” pathways.

**Conclusions:** This work emphasizes a potentially protective role of APOL1 and therefore reverse cholesterol transport in the pathogenesis of GCA. These findings also implicate fibrinolytic and coagulation cascades in GCA susceptibility, highlighting pathways that may be of interest for future pharmaceutical targeting.

**Non-standard Abbreviations and Acronyms:** GCA, giant cell arteritis; MHC, major histocompatibility complex; GWAS, genome-wide association study; SNPs, single nucleotide polymorphisms; PRS, polygenic risk score; pQTL, protein quantitative trait loci; MR, Mendelian randomization; QC, quality control; WTCCC, Wellcome Trust Case Control Consortium; PCA, principal component analysis; IV, instrumental variables; IVW, inverse-variance weighted; PheWAS, phenome-wide association study; GO, gene ontology; FUMA, Functional Mapping and Annotation of Genome-Wide Association Studies; MAGMA, Multi-marker Analysis of GenoMic Annotation; RCT, reverse cholesterol transport.

**Clinical Perspective:** *What is new?:*

- An apoliporotein-L1 polygenic risk score was associated with giant cell arteritis susceptibility, and replicated in an independent dataset.
- Evidence for causality of a protective effect of apolipoprotein-L1 in giant cell arteritis susceptibility was identified using Mendelian randomization.
- Proteins with giant cell arteritis-associated polygenic risk scores were enriched in coagulation-related, fibrinolytic and immune response pathways.

*What are the clinical implications?:*

- Findings from this study indicate a protective role of apolipoprotein-L1 in giant cell arteritis susceptibility, highlighting a potential involvement of reverse cholesterol transport and lipid metabolism in disease pathogenesis.
- Fibrinolytic and coagulation cascades were also implicated in the disease in addition to innate immune response pathways, redrawing attention to the role of thromboinflammation and the need to re-evaluate anti-platelet and anticoagulant therapies, particularly for those with impending visual loss and cranial ischaemic complications.

## Introduction

Giant cell arteritis (GCA) is a medium- and large-vessel vasculitis which overwhelmingly arises in individuals aged ≥ 50 years^1^. GCA is characterized by chronic vascular inflammation (often of cranial arteries) and intimal hyperplasia, resulting in arterial stenoses and downstream ischaemic tissue damage, resulting in clinical manifestations such as sight loss, stroke and scalp necrosis, as well as late aortic dilatation and aneurysm formation^2^.

High incidence rates in populations of Scandinavian descent, not entirely explained by environmental factors, and clustering of familial cases, are indicative of a genetic component to GCA^1, 3^. This is supported by candidate gene studies, which identified genetic associations with GCA in genes of the *major-histocompatibility complex (MHC*)^4^, cytokine genes^5^, and genes with known vascular functions^6^. The largest systematic investigation into the genetic basis of GCA to date is a genome-wide association study (GWAS) on a cohort of 2,134 case and 9,125 control subjects. This study confirmed independent associations in *HLADRB1* and *HLADQA1* in the HLA class II region, alongwith *HLA B* and identified two novel loci related to GCA susceptibility: the *PLG* and *P4HA2* genes, which play important roles in vascular remodelling and angiogenesis, processes previously hypothesised to be involved in the pathogenesis of GCA^7^.

However, given that GCA has a complex, polygenic aetiology, individual genome-wide significant (*P-value*[*P*] ≤ 5 x 10^-8^) variants explain just a small proportion of disease propensity. Previous work investigating complex diseases suggests that genetic risk may be better captured by combining small-effect single nucleotide polymorphisms (SNPs) into a single polygenic risk score (PRS)^8^, which estimates an individual’s propensity to a phenotype by summarising multiple risk alleles carried by an individual, weighted by effect sizes from GWAS summary statistics.

Glucocorticoid monotherapy (usually prednisone) is the principle treatment option for GCA. Research has indicated that whilst glucocorticoid therapy is successful in reducing symptoms of GCA, many patients relapse when tapering glucocorticoid doses or following cessation of treatment. Because of this, patients often remain on low-dose glucocorticoid therapy for many years and experience adverse effects such as osteoporosis, infections, hypertension and gastrointestinal bleeding^9^. Tocilizumab (targeting the interleukin-6 receptor, IL-6R) is a licenced therapy, but access is restricted in many healthcare systems^9^, and not all patients can tolerate this therapy^10^. Thus, there is ongoing unmet need for novel targeted therapies and improved biomarkers for risk stratification to induce and maintain remission of GCA.

It has been demonstrated that a drug with additional genetic support is twice as likely to proceed from phase I clinical trials to approval^11, 12^. Therefore, using genetic data to guide drug discovery could reduce rates of failure in clinical development caused by poor drug efficacy, particularly when genetic data are enriched with other types of ‘omics information, such as proteomics^13^. By doing so, it may be possible to identify proteins implicated in the pathogenesis of GCA and provide a basis for pharmaceutical targeting and biomarker identification.

For example, recent developments in high-throughput technologies and the emergence of genome-wide methodology have allowed groups to perform large-scale protein quantitative trait loci (pQTL) analyses and subsequent causality estimation with Mendelian randomisation (MR)^13, 14^. Such work has identified inherited variation in protein levels which could provide the link between germline genetic variants and disease phenotypes, given that proteins are often the molecules directly implicated in pathogenic cascades.

In this study, we constructed PRS for several blood secretome proteins and tested these for association with GCA. We used MR to test for causality of these proteins and suggest novel biomarkers for drug repurposing or patient stratification.

## Methods

### Protein selection

Circulating plasma proteins with publicly available summary statistics, generated from GWAS investigating inter-individual variation in protein levels, were selected for study in this work^13^. To prioritise the proteins investigated and minimise multiple testing, two filters were applied to the selection of plasma proteins for inclusion in the study. Firstly, 730 proteins deemed part of the “human blood secretome” were selected, based on prior knowledge that circulating proteins represent useful druggable targets and because their primary physiological action is in circulating form^15^. Secondly, proteins with low estimated levels of heritability (*R^2^*≤lowest quartile), therefore underpowered to demonstrate strong genetic associations with GCA-risk, were removed from the study. Following these filters, 169 proteins remained for subsequent analysis (**Supplementary Methods**).

Quality control of giant cell arteritis datasets

Cases from UK GCA consortium (*N*=1,858) and controls from Wellcome Trust Case-Control Consortium (WTCCC; *N*=3,748), were used to develop PRS for blood secretome proteins in this work. Information regarding the quality control (QC) of both GCA and Wellcome Trust Case Control Consortium (WTCCC) genotype data may be found in **Supplementary Methods**.

Briefly, genotyping was performed using the Illumina “Infinium HumanCore Beadchip” and “Infinium Global Screening” arrays (UK GCA cases) and Illumina 1.2M custom chip (WTCCC controls, late summer 2009 release). Sample and variant QC were executed in PLINK v1.07^16^, including the use of principal component analysis (PCA) to account for population stratification (**Supplementary Methods**).

This cohort was divided into two subsets. The first comprised 729 GCA cases and 2,619 WTCCC controls (from the 1958 British Birth Cohort), and was used to optimize protein PRS and test for their association with GCA susceptibility.

The second subset was used for PRS replication in this work, formed from a cohort of 1,129 UK GCA consortium cases and 2,654 WTCCC controls (from the UK Blood Service control group).

### Polygenic risk scoring

The PRSice v2.3.3^17^ software was used for developing protein PRS and testing for associations with GCA. PRS were constructed via the “clumping and thresholding” approach, using effect sizes and *P-values* from GWAS of protein levels^13^, and tested for association with GCA susceptibility in the primary sub-cohort using logistic regression (*GCA ∼ PRS + PC1-10*; **Supplementary Methods**). To account for population stratification, the top 10 principal components (PCs) from PCA were included as covariates. The “optimal” PRS was defined as that PRS with the greatest Nagelkerke^18^ *R^2^*value in logistic regression.

To account for multiple testing, the significance threshold was adjusted for the number of proteins tested using Bonferroni correction. For the 169 proteins assessed by PRS analysis, the corrected *P-value* threshold was 0.05/169=2.96x10^-4^.

Details of additional sensitivity analyses, including testing of associated PRS for different case definitions, and different GCA severity outcomes, may be found in **Supplementary Methods** (**Supplementary Table 1**).

Replication was performed using the secondary sub-cohort. SNPs and weights from GCA-associated protein PRS were used to calculate PRS in an independent dataset with PRSice v2.3.3^17^, and these PRS were tested for association with GCA in logistic regression, using the top 10 PCs from PCA as covariates (*GCA ∼ PRS + PC1-10*). PRS were calculated using SNPs from data unfiltered for imputation quality. This was in order to capture as many SNPs as possible from PRS developed in the primary analyses, thereby reproducing PRS as accurately as possible. 99.98% of imputed SNPs in these PRS had an imputation quality R^2^>0.3, and no SNP had an R^2^<0.04.

### Testing polygenic risk scores for association with off-target proteins

Use of SomaLogic aptamers in protein quantification holds potential for cross-reactivity with off-target proteins. Therefore, where possible, PRS with statistically significant associations with GCA were tested for association with proteins found in complex with target proteins (identified using a literature search), to assess whether PRS were predictive of these protein levels instead of intended targets. Protein quantification data was obtained from UK Biobank^19^ and tested for association with the relevant PRS using linear regression in up to 350,883 white European UK Biobank participants, adjusting for the top 10 PCs from PCA as covariates. Details of UK Biobank QC and PCA may be found in **Supplementary Methods**.

### Mendelian randomization

Proteins with GCA susceptibility-associated PRS were further evaluated using MR, to assess whether their associations with GCA represent a causal relationship. MR is a statistical technique which uses genetic variants as instrumental variables (IV) to assess whether ‘exposures’ (here, proteins) are causally associated with an outcome (here, GCA).

Using summary statistics from respective GWAS of plasma protein levels^13^, three genetic scores were constructed as IVs (following the pruning of SNPs with linkage disequilibrium *R^2^*>0.1).

IVs were then used to tested for an association between the protein and GCA susceptibility in MR, using summary stastistics from a GCA GWAS. The three scores for each tested protein used a different *P-value* threshold to filter SNPs for inclusion in the IV. These included: a “liberal” score (*P-value* threshold as defined by the optimal PRS in the clumping and thresholding approach), an “intermediate” score (*P-value* threshold≤5 x 10^-5^), and a “conservative” score (*P-value* threshold≤5 x 10^-8^).

Two-sample MR was performed using the inverse-variance weighted (IVW) method, and the measure of variance used was standard error (SE). Sensitivity analyses (including Cochran’s heterogeneity test, MR-Egger intercept test, and weighted median and mode MR methods) were performed to rule out potential horizontal pleiotropy, which could invalidate use of the genetic instruments^14^. If IVs yielded evidence for potential pleiotropy in sensitivity analyses, SNPs from that IV were further investigated using a phenome-wide association study (PheWAS) approach (**Supplementary Methods**).

### Protein-protein interaction network analysis

Proteins with associated PRS were tested for protein-protein interaction enrichment using StringDB v12.0^20^. StringDB utilizes multiple sources of evidence (e.g. experimental, coexpression, and text mining) as priors to assess the probability of two proteins being functional interactors. Enrichment of gene ontology (GO) processes for proteins in this network were also tested^21^.

### Pathway analyses

The Functional Mapping and Annotation of Genome-Wide Association Studies (FUMA) v1.4.0 tool^22^ was used to perform pathway analysis on protein PRS associated with GCA. FUMA is a package which combines several *in-silico* tools (including the Multi-marker Analysis of GenoMic Annotation (MAGMA) gene-based test^23^) to provide functional interpretation of SNPs in PRS.

## Results

### Polygenic risk scoring

We developed PRS for 169 proteins/protein complexes of the human blood secretome. Of these PRS, 12 reached *P*<0.05 prior to multiple testing adjustment (**Figure 1**), whilst one protein, apolipoprotein-L1 (APOL1), remained significant at *P*=2.96x10^-4^ following Bonferroni correction (**Table 1**).

**Figure 1.**
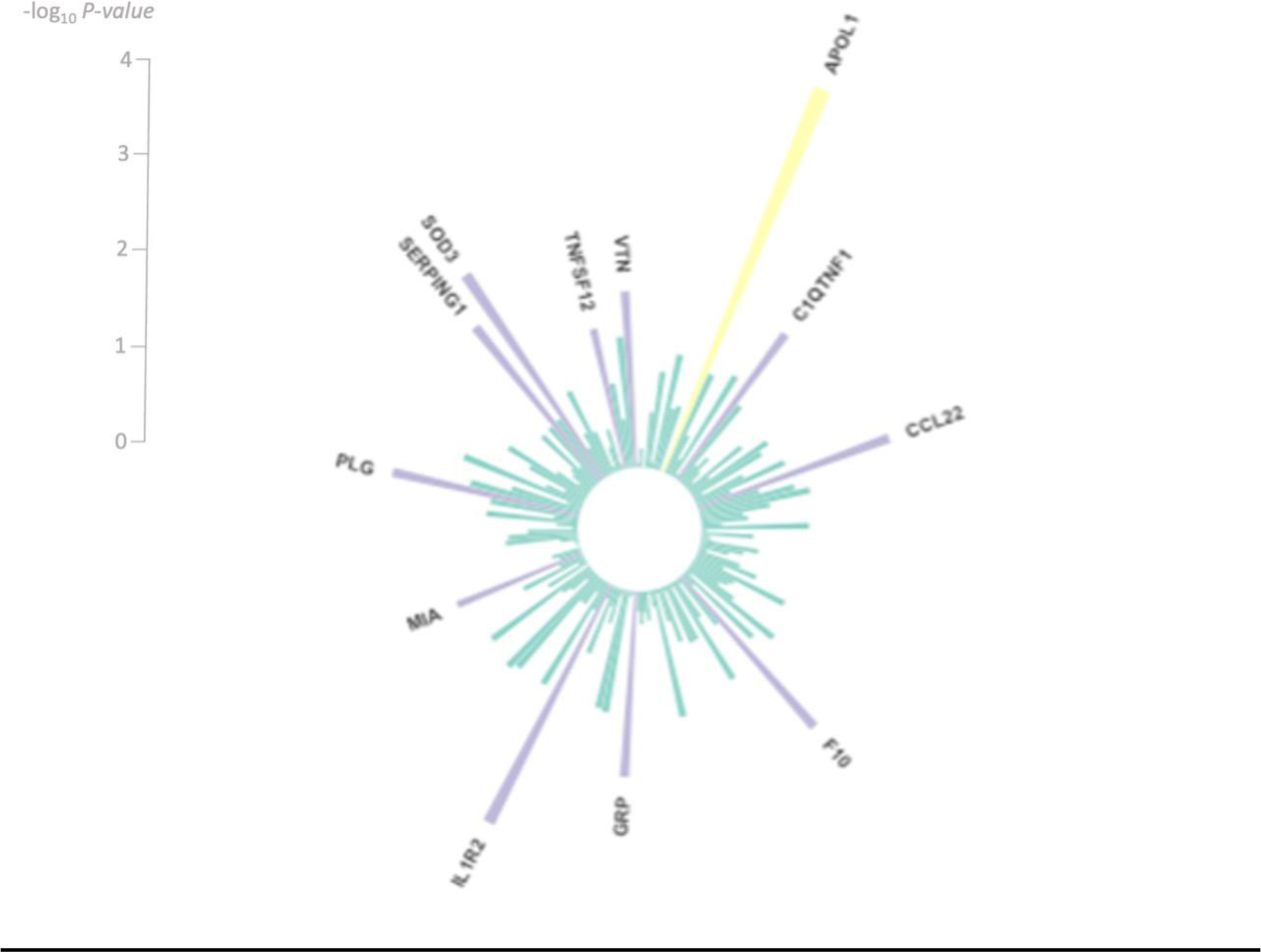
Circos plot of the -log_10_ *P-values* for association between the “best fit” polygenic risk scores (generated using protein quantitiative trait loci (pQTL) summary statistics for each protein) and giant cell arteritis (GCA) case-control status. A significant association indicates that a single polygenic risk score for abundance of a particular protein also predicted GCA risk; the *P-value* (*P*) of this association is represented by the length of bar on the plot. These scores indicate shared genetic aetiology between the traits and are suggestive of a causal link between the protein levels and disease outcome. Blue bars represent associations not reaching the *P* ≤ 0.05 threshold. Purple bars represent associations significant at *P*≤0.05 but which did not pass the Bonferroni-corrected *P*≤2.96x10^-4^ threshold. Yellow bars represent associations at the *P*≤2.96x10^-4^ Bonferroni- corrected threshold.

**Table 1.**
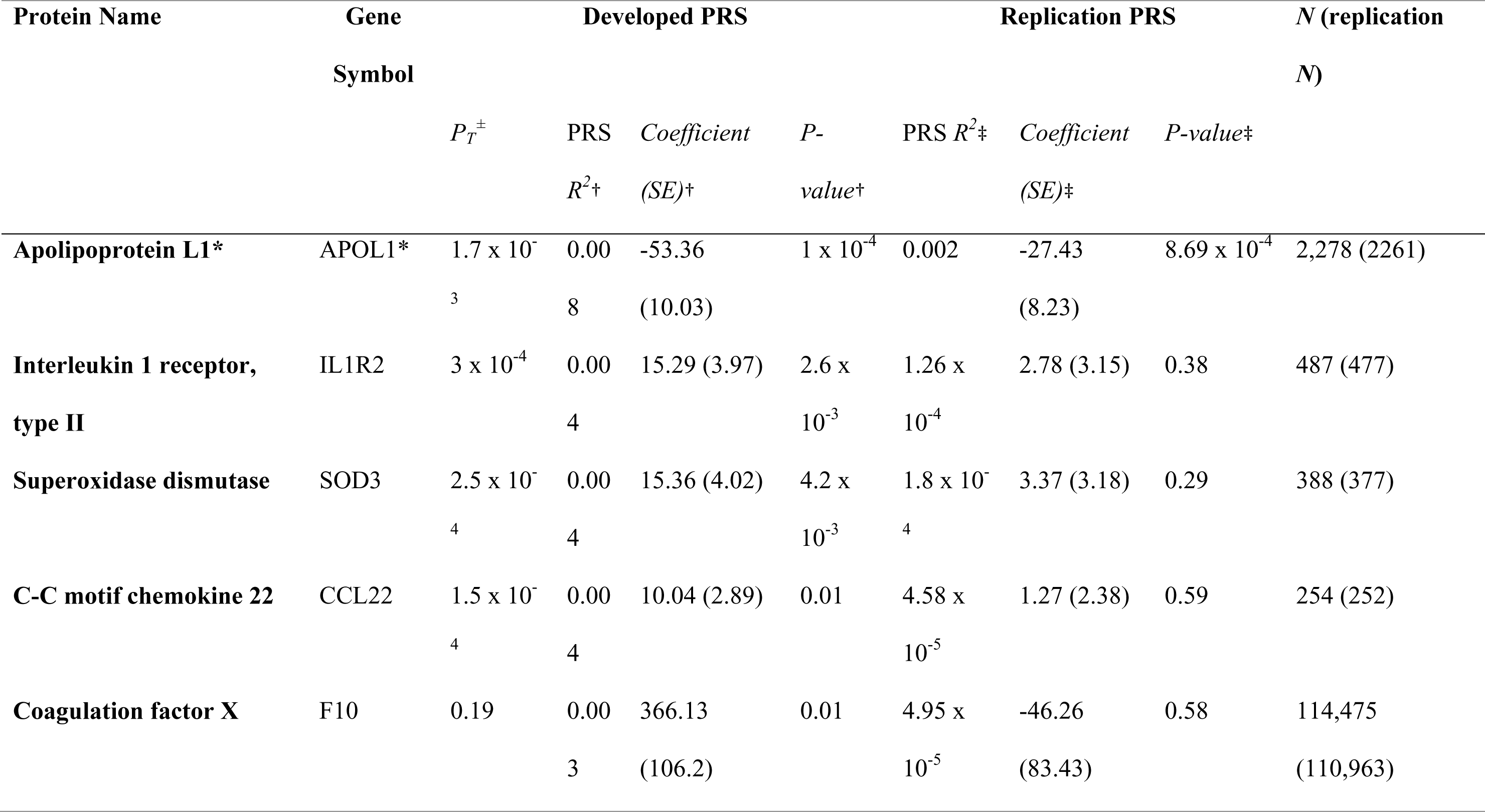
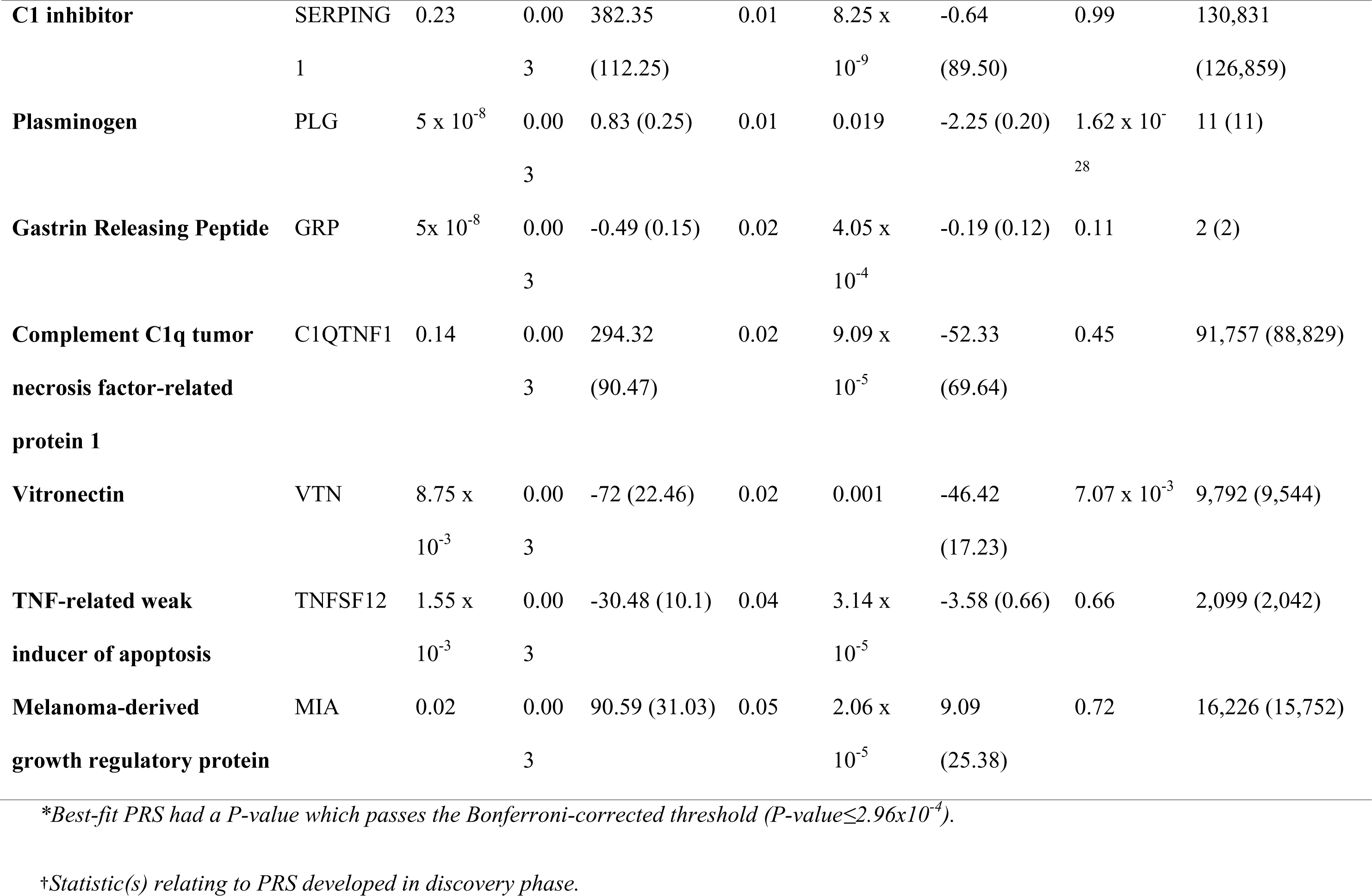
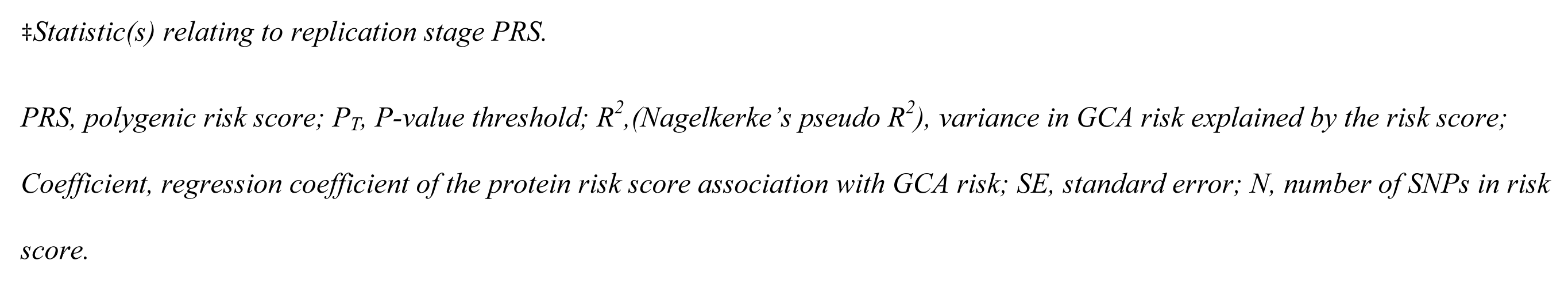
Best-fit polygenic risk scores for the 12 proteins with a PRS association *P-value* < 0.05.

The APOL1 PRS had a statistically significant association with GCA susceptibility (*P*=1x10^-4^), following Bonferroni correction. This PRS consisted of 2,278 SNPs distributed across the genome, two of which (rs117674301 and rs117850190) were classed as *cis*-pQTL, whilst most had *trans* effects on APOL1 levels. The PRS had a negative coefficient (*coefficient[SE]*=- 53.36[10.03]), indicating that a PRS which increases levels of APOL1 is associated with reduced risk of GCA (**Figure 2**). The variance in GCA susceptibility explained by the risk score is estimated at approximately 0.8% (Nagelkerke’s *R^2^*=0.008). Results of high-resolution PRS for APOL1, testing the APOL1 PRS in different case definitions, and testing for association between APOL1 and secondary outcomes may be found in **Supplementary Results** (**Supplementary Figures 1-2; Supplementary Table 2**).

**Figure 2.**
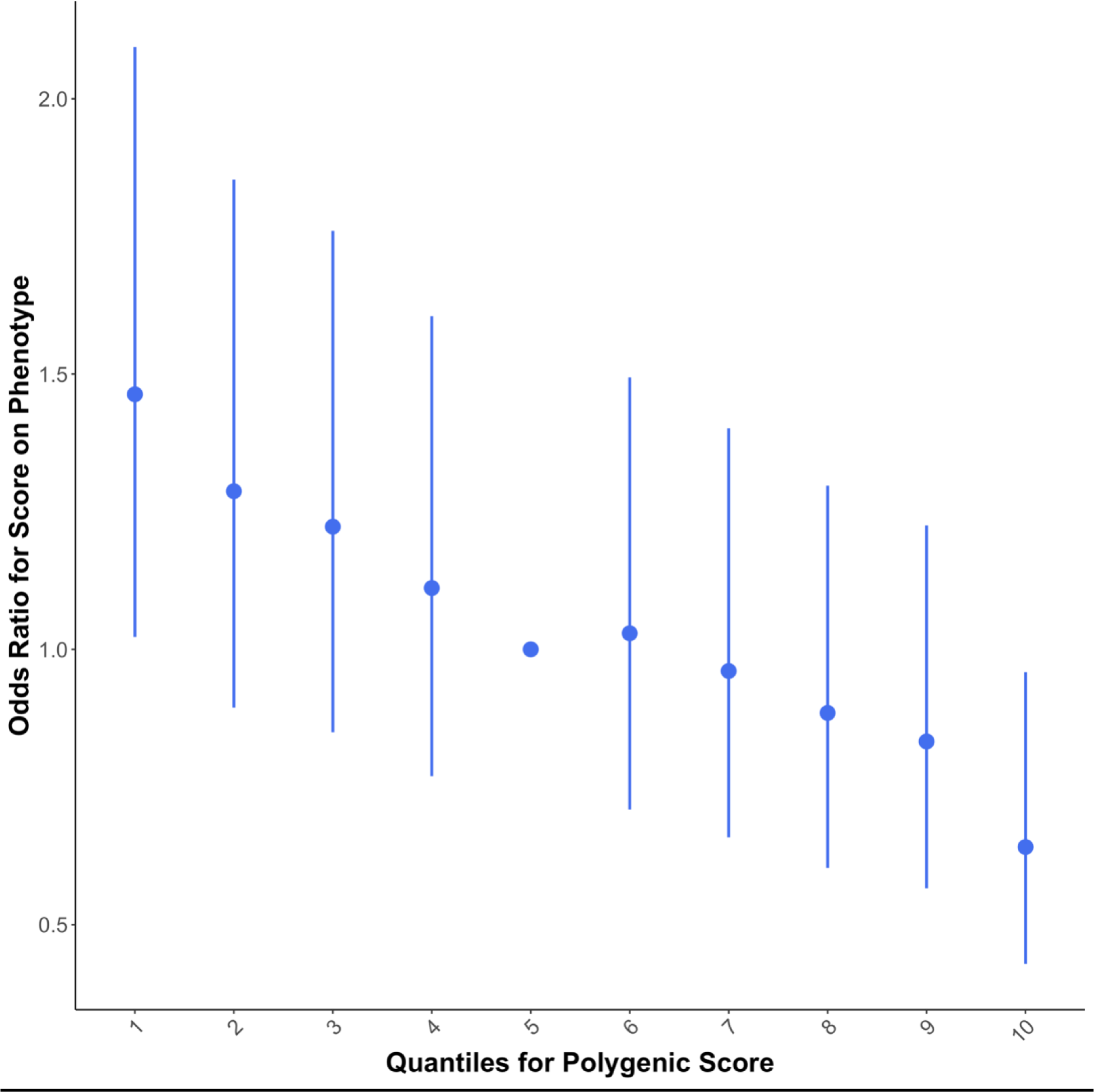
Quantile plot demonstrating the effect of increasing the optimal polygenic risk score (PRS) from Apolipoprotein L1 (APOL1) protein quantitative trait loci (pQTL) on giant cell arteritis (GCA) risk. Samples were assigned to a non-overlapping decile based on their PRS, and logistic regression was performed to generate odds ratios for the association between the APOL1 PRS decile and GCA risk. The fifth quantile is the reference and error bars around points represent 95% confidence intervals. Increasing the APOL1 PRS value is associated with increased plasma levels of APOL1 and a decreased risk of GCA.

PRS associated with GCA susceptibility passing the *P-value*<0.05 are summarised in **Supplementary Results** (**Table 1**). Briefly, these associations included PRS for the superoxidase dismutase protein (*coefficient [SE]*=15.36 [4.02]; *P*=4x10^-3^), C1 inhibitor protein (*coefficient[SE]*=382.35[112.25]; *P*=0.01), and plasminogen protein (*coefficient[SE]*=0.83[0.25]; *P-value*=0.01).

Replication of the APOL1 PRS association was performed in an independent GCA dataset comprising 1,129 cases and 2,654 controls. Of the 2,278 SNPs in the initial PRS, 2,261 were present in the second dataset (**Table 1**). Following adjustment for 10 PCs, the APOL1 PRS had a statistically significant association with GCA with a negative direction of effect (*coefficient[SE]*=-27.43[8.23], *P*=8.69x10^-4^), consistent with that found for the association in the primary cohort.

Furthermore, 11 other protein PRS with *P-value*<0.05 were tested for replication, revealing two associations: the plasminogen PRS (*R^2^*=0.019; *coefficient*[*SE*]=-2.25[0.20]; *P* =1.62x10^-28^) and the vitronectin PRS (*R^2^*=0.001; *coefficient*[*SE*]=-46.42[17.23]; *P*=7.07x10^-3^).

### Testing polygenic risk scores for association with off-target proteins

To determine whether the APOL1 PRS is associated with off-target proteins, it was tested for association with APOA1 blood plasma levels (UK Biobank nuclear magnetic resonance measurements) in a linear regression utilising 350,883 UK Biobank subjects. No association was found between the APOL1 PRS and APOA1 (*coefficient*=-0.34; *SE*=0.65; *P*=0.60), suggesting that the PRS is not predictive of APOA1 levels.

### Mendelian randomization

Once effect and non-effect alleles were harmonised between the exposure (APOL1) and outcome (GCA susceptibility) data, MR analysis was performed using each of the three risk scores constructed for use as APOL1 IVs. Using the IVW method of MR, the APOL1 protein was estimated by all risk scores to have a statistically significant, causal effect on GCA susceptibility (**Supplementary Table 3; Supplementary Figure 3**). The direction of effect was negative in each instance (liberal *beta*[*SE*]=-0.093[0.02]; intermediate *beta*[*SE*]=-0.131[0.05]; conservative *beta*[*SE*]=-0.22[0.1]), and was supported by the weighted median and weighted mode methods of MR. Details of MR sensitivity analyses may be found in **Supplementary Results (Supplementary Tables 4–7)**. Briefly, results of the MR-Egger intercept test lacked statistical significance for any of the risk scores, and following the repetition of MR with three potentially pleiotropic SNPs removed from the score, the causal effect of the APOL1 protein on GCA susceptibility remained statistically significant for each of the risk scores. This indicates a lack of evidence to suggest that pleiotropic SNPs are confounding the association observed here.

### Protein-protein interaction network analysis

To understand relationships between proteins with GCA susceptibility-associated PRS, we performed protein-protein interaction [PPI] network analysis (**Figure 3**). Significantly more interactions were observed across the 12-protein network than expected by chance (nodes *N*=12, edges *N*=6, expected edges *N*=1, PPI enrichment *P*=1.61x10^-4^). The most significantly enriched GO processes for proteins in this network included “negative regulation fibrinolysis” (pathway protein [PP] *N*=13, network protein [NP] *N*=2, *false discovery rate*[*FDR*]=0.04), and “negative regulation of blood coagulation” (PP *N*=46, NP *N*=4, *FDR*=2.8x10^-4^).

**Figure 3.**
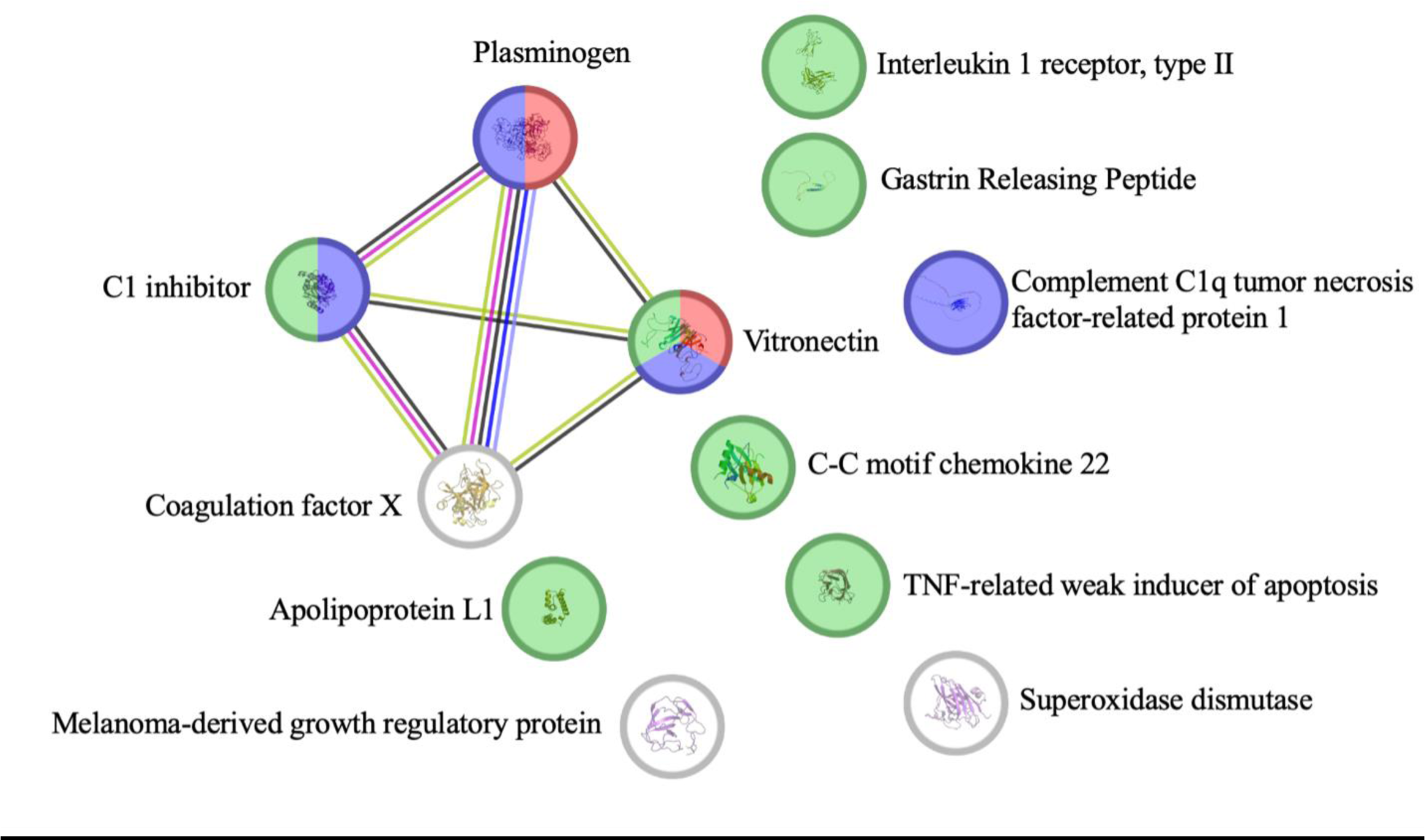
StringDB network analysis using proteins (nodes) whose PRS had “borderline” significant (*P-value*≤0.05) associations with GCA susceptibility. Nodes are coloured according to presence in enriched gene ontology biological pathways, including: “negative regulation of fibrinolysis” (red), “negative regulation of blood coagulation” (blue) and “immune response” (green). Edges of this network represent protein-protein links found by StringDB via textmining (green), coexpression (black), protein homology (light blue), gene co-occurrence (navy), and experimental (pink) evidence.

### Pathway analyses

Pathway analyses were performed on the 2,278 SNP APOL1 PRS. Using the MAGMA gene- based test^23^, input SNPs were assigned to protein-coding genes (*N*=19,054), revealing 14 genes significantly enriched for SNPs in the APOL1 PRS at the Bonferroni corrected *P-value* 0.05/19,054=2.62x10^-6^ (**Table 2**; **Figure 4**), including: *DNMBP* (*z-stat*=8.34; *P*=3.81x10^-17^), *LRRC15* (*z-stat*=7.75; *P*=4.77x10^-15^), *CPN1* (*z-stat*=6.11; *P*=5 x 10^-10^), and *CPN2* (*z-stat*=6.11; *P*=5x10^-10^).

**Figure 4.**
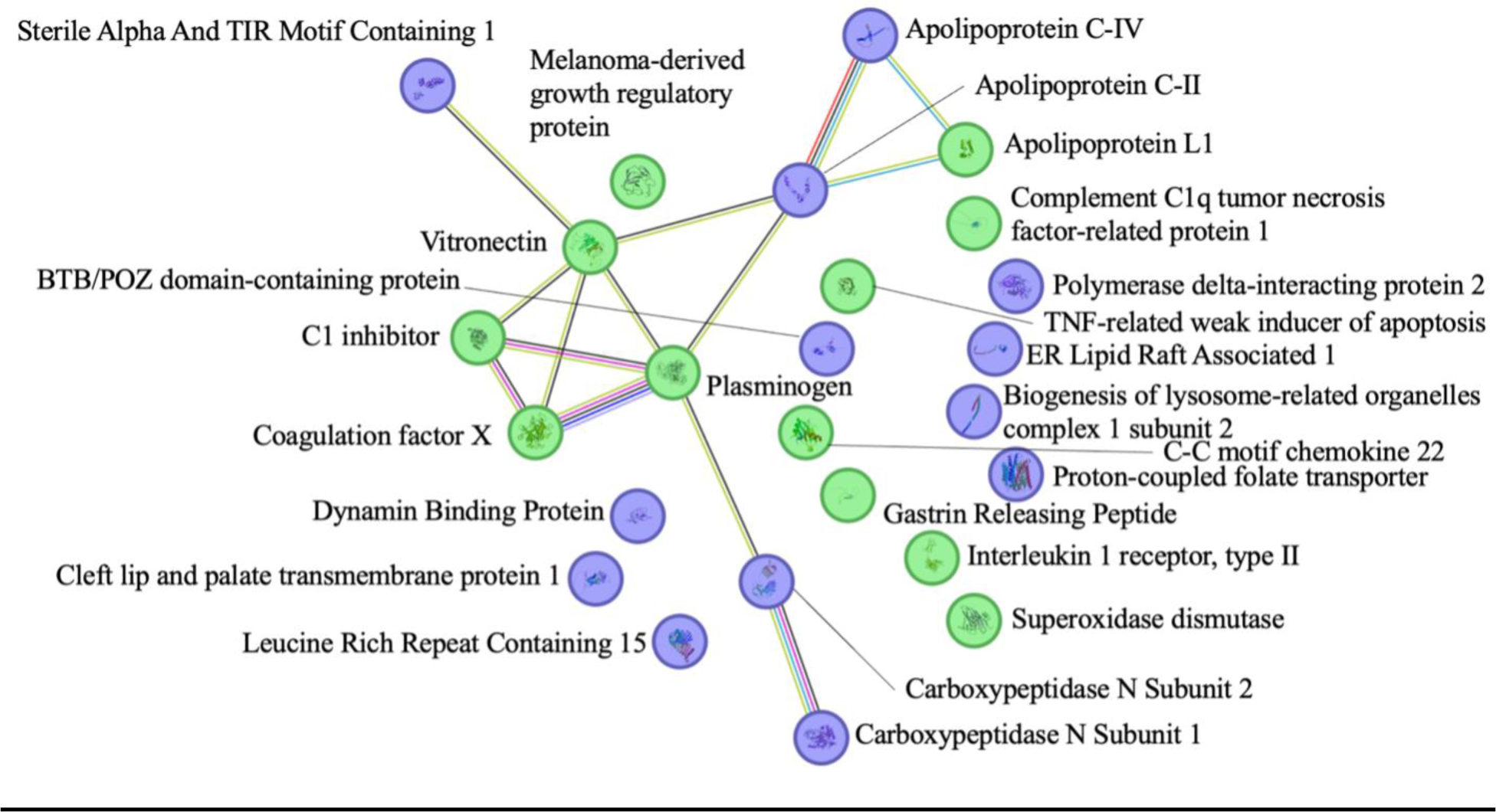
StringDB network analysis using proteins (nodes) whose PRS had “borderline” significant (*P-value*≤0.05) associations with GCA susceptibility (green nodes), or whose respective genes were enriched for SNPs in the APOL1 PRS (blue nodes). Edges of this network represent protein-protein links found by StringDB via textmining (green), coexpression (black), protein homology (light blue), gene co-occurrence (navy), experimental (pink), and gene fusion (red) evidence.

**Table 2.** Results of the Multi-marker Analysis of GenoMic Annotation (MAGMA) gene- based test, performed using SNPs of the apolipoprotein L1 PRS.

| Gene Symbol | Encoded Protein | Chr | SNPs *N* | *Z* | *P* |
| --- | --- | --- | --- | --- | --- |
| *DNMBP* | Dynamin Binding Protein | 10 | 590 | 8.3369 | 3.81 x 10<sup>-17</sup> |
| *LRRC15* | Leucine Rich Repeat<br>Containing 15 | 3 | 113 | 7.7451 | 4.77 x 10<sup>-15</sup> |
| *CPN2* | Carboxypeptidase N Subunit 2 | 3 | 80 | 6.1094 | 5.00 x 10<sup>-10</sup> |
| *CPN1* | Carboxypeptidase N Subunit 1 | 10 | 154 | 6.1094 | 5.00 x 10<sup>-10</sup> |
| *CLPTM1* | Cleft lip and palate<br>transmembrane protein 1 | 19 | 140 | 6.1038 | 5.18 x 10<sup>-10</sup> |
| *SLC46A1* | Proton-coupled folate<br>transporter | 17 | 20 | 5.3177 | 5.26 x 10<sup>-8</sup> |
| *BLOC1S2* | Biogenesis of lysosome-related<br>organelles complex 1 subunit 2 | 10 | 48 | 5.2006 | 9.93 x 10<sup>-8</sup> |
| *APOC4* | Apolipoprotein C-IV | 19 | 32 | 4.9299 | 4.11 x 10<sup>-7</sup> |
| *APOC4-* | Apolipoprotein C- | 19 | 32 | 4.9299 | 4.11 x 10<sup>-7</sup> |
| *APOC2* | IV/Apolipoprotein C-II |  |  |  |  |
| *POLDIP2* | Polymerase delta-interacting<br>protein 2 | 17 | 17 | 4.769 | 9.26 x 10<sup>-7</sup> |
| *TNFAIP1* | BTB/POZ domain-containing<br>protein | 17 | 21 | 4.7196 | 1.18 x 10<sup>-6</sup> |
| *CTB-96E2.2* | - | 17 | 14 | 4.6054 | $2.06 \times 10^{-6}$ |
| *SARM1* | Sterile Alpha And TIR Motif<br>Containing 1 | 17 | 103 | 4.5917 | $2.20 \times 10^{-6}$ |
| *ERLIN1* | ER Lipid Raft Associated 1 | 10 | 100 | 4.5843 | $2.28 \times 10^{-6}$ |

*SNP, single nucleotide polymorphism; PRS, polygenic risk score; Chr, chromosome; N, number; Z, z-stat; P, P-value.*

When proteins endoded by these genes were added to the protein-protein interaction network analysis, significantly more interactions were observed across the 25 proteins than would be expected by chance (nodes *N*=25, edges *N*=14, expected edges *N*=2, PPI enrichment *P- value*=1.69x10^-9^). The most significantly enriched GO processes for proteins in this network included “negative regulation of blood coagulation” (PP *N*=46, NP *N*=4, *FDR*=7x10^-3^), and “immune response” (PP *N*=1,321, NP *N*=9, *FDR*=0.04).

## Discussion

Here, we performed a systematic evaluation testing genetically-determined levels of circulating proteins against GCA susceptibility. We found evidence for an association between a 2,278 SNP APOL1 PRS and GCA, which replicated in an independent cohort. Further evaluation with MR revealed evidence for causal effect on disease risk, with genetic tendency to higher levels of APOL1 plasma protein associated with reduced risk of GCA. These results are suggestive of a potentially protective role for circulating APOL1 protein in GCA and highlight a shared genetic aetiology of the protein and GCA.

This association emphasises the potential benefits of integrating genomic and proteomic data when searching for novel pathogenic mechansisms, and hence potential therapeutic targets, in disease. Pairing genetic information with protein data allows not only the identification of an association between proteins and disease, but enables the discovery of common biological pathways influencing both protein levels and disease pathogenesis, providing more research avenues for the identification of novel therapeutic targets and risk stratification biomarkers.

APOL1 arose from a gene duplication event approximately 30 million years ago and is present in some higher primate species and humans, but not other mammals^24^. It is an innate immune effector which integrates into protein complexes and circulates in blood plasma bound to high density lipoprotein (HDL) particles or complexed to IgM^25^. It is thought to have roles in protection against pathogens, inflammation, lipid binding/transport (including cardiolipins), cholesterol metabolism, and cardiovascular disease^25, 26^.

Much of our biological knowledge of APOL1 is derived from studies of recessive gain of function *APOL1* variants that emerged in Sub-Saharan Africa, which conferred protection from pathogenic trypanosomes. Approximately 13% of African Americans have two copies of risk alleles, which explains most of the excess risk of non-diabetic kidney disease in individuals of African descent, particularly hypertensive end-stage kidney disease and focal segmental glomerulosclerosis, but also lupus-related nephropathy. Poor renal transplant outcomes observed when the donor tissue, rather than recipient, has the *APOL1* risk haplotypes, suggests the poor renal outcomes are likely to be due to a tissue phenotype rather than circulating APOL1 levels^24^.

Although the APOL1 protein is found in kidney tissue, the majority of APOL1 in humans is found circulating in serum or located in vascular tissue^27^. Previous work has suggested that plasma APOL1 levels are not associated with kidney function, but do correlate with fasting lipids, suggesting that the source of APOL1 is unlikely to be from kidney tissue ^28, 29^. Proteomics approaches have demonstrated that APOL1 circulates as part of two specialized complexes which form a minor HDL subfraction. In the first, APOL1 is bound to cholesterol, cholesterol esters, phospholipids, haptoglobin-related protein (HPR), haemoglobin and another apolipoprotein, APOA1, forming a complex termed trypanosome lytic factor-1 (TLF1). In the second, APOL1 is found as part of a complex again containing APOA1 and HPR, but also immunoglobulin M and fibronectin, termed TLF2^25^.

Interestingly, TLF complexes and APOL1 have proposed functions in HDL metabolism, and are linked to susceptibility to CVDs^25^. As previously established, APOL1 is a minor protein component of HDL and is expressed in human vascular cells. Important roles of HDL are thought to be reverse cholesterol transport (RCT) from peripheral tissues to the liver, and promotion of nitric oxide production by endothelial cells, resulting in anti-apoptotic/anti- inflammatory effects and a reduction in atherosclerotic risk^30^. Previous work has found that African ancestry risk variants APOL1-G1 and –G2 (which are thought to negatively impact APOL1 function and are found in low frequencies in European populations with minor allele frequency 0.000242 and 0.00013, respectively^31, 32^), result in impaired RCT via reduction in expression of cholesterol efflux transporters^33^; thus providing evidence for the role of APOL1 in RCT. It has been proposed that with impaired RCT, lipid accumulation, macrophage transformation into foam cells and release of inflammatory factors occur, creating a pro- inflammatory state which could lead to increased ischaemic risk.

The impact of APOL1 levels on impaired RCT is supported by evidence that found low serum levels of APOL1, along with low HDL-bound APOL1 levels (affecting HDL composition, but not total HDL levels) in subjects with familial hyperlipidemia, suggesting that low volumes of APOL1 may reduce RCT and result in increased cholesterol/lipid levels in individuals. Additionally, the study found that low APOL1 levels were predictive of ischaemic cardiac

events, and that APOL1/HDL-cholesterol ratios were predictive of post-event survival rates^34^. It is therefore possible that impaired cholesterol transport and subsequent lipid accumulation, atherosclerosis and inflammatory mediator release could represent a mechanism through which low heritable APOL1 levels may contribute to the increased vascular inflammation found in GCA patients.

Because APOL1 is almost exclusively found bound to complexes with the APOA1 protein, it was important to ensure that the PRS found to be associated with GCA in this work was predictive of levels of APOL1 in blood plasma, and not APOA1. To test this, the APOL1 PRS developed in this work was tested for association with APOA1 levels in UK Biobank data. No association was found between the PRS and APOA1 levels, indicating that the PRS is likely predictive of APOL1 levels, and not APOA1, in the plasma.

In addition to the APOL1 PRS, a small number of PRS had associations at the *p* ≤ 0.05 level, but did not yield statistical strength to pass the multiple testing threshold. Of these results, two PRS replicated in an independent cohort: vitronectin PRS and plasminogen PRS, providing evidence that these proteins may also be implicated in GCA susceptibility, supporting previous findings from GWAS^35^. Protein-protein interaction network analysis of these proteins using StringDB^20^ revealed biological pathways enriched for proteins in the network, including “negative regulation of fibrinolysis”, “negative regulation of blood coagulation” and “immune response” pathways.

When proteins encoded by genes enriched for SNPs in the APOL1 PRS were added to the protein-protein interaction network, both the “negative regulation of blood coagulation” and “immune response” pathways remained enriched, and a 10-protein network was formed, including vitronectin, plasminogen, C1 inhibitor, coagulation factor X and multiple apolipoproteins. Notably, this network links apolipoprotein L1 to the vitronectin-plasminogen

network through apolipoprotein C2, a protein which plays a known role in lipoprotein metabolism and has recently been implicated in Takayasu arteritis^36^. These findings may suggest a link between both lipid metabolism and coagulation cascades, and the development of GCA, and highlight a number of new potential biomarkers for future study. Indeed, given prior speculation over the role of anti-coagulants in reducing risk of cranial ischaemic complications in GCA^37^, it is prudent that these pathways are further interrogated using functional and multi-omics studies to untangle the relationship between fibrinolytic/coagulation cascades and GCA progression.

### Limitations

Findings from this study must be interpreted in light of some limitations. This work’s primary drawback concerns its limited sample size. A small cohort can restrict power, meaning that variants of small effect may be missed (although this was combated using a PRS approach). Furthermore, two filters were applied during selection of plasma proteins, in order to reduce the number of tested hypotheses. However, by limiting the number of proteins studied, molecules important in the pathogenesis of GCA may have been overlooked. For example, only proteins present in the blood secretome were retained for analysis, and some molecules implicated in GCA (such as JAK2)^38^ were excluded for this reason. It may therefore be beneficial to repeat analyses using a wider range of proteins in future studies utilising larger cohorts.

A further limitation of the work concerns the replication of the APOL1 PRS in an independent cohort. Whilst the PRS was developed using genetic data filtered for imputation quality, the second cohort in which it was calculated did not have an imputation quality threshold applied. This was in order to accurately replicate the developed PRS. However, the absence of this threshold could have resulted in the inclusion of poorly imputed SNPs in the PRS, adding noise to the PRS and potentially impacting power of the PRS-GCA association. Future work should aim to disentangle the impact of these SNPs on APOL1 PRS performance and replicate these findings in a third, independent cohort.

Finally, whilst MR is a useful statistical concept for generating causal estimates, it is dependent on a number of assumptions. Previous work has indicated that whilst large risk scores can improve MR results by increasing power, the validity of these risk scores may be reduced if the SNPs are not true predictors of the exposure variable^14^. Traditionally, genome-wide significant SNPs at the gene locus of the exposure are used to ascertain this; however, the purpose of this study was to identify genome-wide pQTL that affect protein levels, so sole use of *cis*-pQTL would be inappropriate. Instead, the combined power of *cis*- and *trans*-pQTL in a PRS should provide sufficient statistical and biological justification for their use as IVs. Furthermore, two additional risk scores, consisting solely of variants with greater evidence for association with APOL1, were used to confirm results found by the initial analyses. Because little is known about the aetiology of GCA, the second and third assumptions, which regard alternate pathways from SNPs to GCA, were difficult to assess objectively. For this reason, MR was used primarily to support PRS findings, and results from these analyses should not be interpreted alone.

## Conclusion

In this study interrogating human secretome proteins using a PRS approach, a 2,278 SNP APOL1 PRS was associated with GCA susceptibility, with further evidence for causality using MR. The findings from this work has revealed possible insights into the pathogenesis of GCA, and the APOL1 protein may represent a future biomarker therapeutic targeting. For the first time, these findings highlight the potential roles of RCT and the trypanosome lytic factors in GCA pathogenesis, and demonstrate the value of repurposing publicly available genetic data in discovery work.

## Data Availability

Data access requests should be directed to the corresponding author. We are unable to share UK Biobank data but have included the variable names so that the study can be reproduced. Generated PRS are available on request.

## Acknowledgements

We thank all patients who participated in this study. All UK GCA Consortium group members contributed to patient and data collection and were integral to the delivery of this project. A list of group members can be found in Appendix 1, available online.

## Ethics approval

A favourable ethical opinion for the UK GCA Consortium (UK GCA) study was granted by the Yorkshire and the Humber Leeds West Research Ethics Committee (05/Q1108/28).

## SupplementaryMaterial

### SupplementaryMethods

#### Quality control of giant cell arteritis datasets

A case-control cohort comprised of samples from the UK giant cell arteritis (GCA) consortium (*N*=1,858) and the Wellcome Trust Case-Control Consortium (WTCCC; *N*=3,748) was used to optimize polygenic risk scores (PRS) in this work. Informed written consent was gained from all participants (Yorkshire and the Humber Leeds West Research Ethics Committee [05/Q1108/28]), including 1,858 cases with a confirmed clinical (consultant rheumatologist or ophthalmologist) diagnosis of GCA (i.e. biopsy confirmed, positive imaging or adequate symptomatic characteristics to allow for a sound clinical diagnosis). Of these individuals, 963/1,386 (69.48%) had a confirmed diagnosis (temporal artery biopsy[TAB] or imaging, including: magnetic resonance angiography [MRA], positron emission computed tomography with 2-deoxy-2-fluorine-18-fluoro-D-glucose [^18^FDG-PET/CT], ultrasound scan [USS], computed tomography [CT], and CT angiography [CTA]). Additionally, 1,287/1,379 cases (93.32%) fulfilled imputed American College of Rheumatology classification criteria^9^, including at least three of: new headache, age at disease onset ≥ 50 years, abnormal TAB, temporal artery abnormality, and heightened ESR (≥ 50 mm/hr). In this work, ESR was imputed from CRP, due to a lack of routine ESR measurements at several UK NHS sites at the time of data collection.

For cases with both ESR and CRP available (*N*=1001), missing variable percentile analysis was performed, and it was found that for an ESR ≥ 50mm/hr, the equivalent CRP measurement was 36mg/L (9th percentile). Therefore, for cases with missing ESR values, CRP measurements ≥ 36 mg/L were used to impute ESR ≥ 50mm/hr (elevated ESR).

Three ischaemic phenotypes were defined in this cohort. Cranial ischaemic complications in GCA were defined as: non-ocular cranial complications such as tongue necrosis, scalp necrosis, or cerebrovascular accident at presentation (secondary to GCA); and ocular complications such as cranial nerve palsy (III, IV, or V), branch retinal artery occlusion, central retinal artery occlusion, cilioretinal artery occlusion, posterior ischaemic optic neuropathy, Anterior ischaemic optic neuropathy, irreversible visual loss, irreversible visual field defect, irreversible ocular motility, irreversible diplopia, or relative afferent pupillary defect. Transient cranial ischaemic manifestations were defined as: non-ocular cranial ischaemic features such as transient ischaemic attack at presentation (secondary to GCA), tongue claudication, or jaw claudication; and ocular ischaemic features such as transient vision loss, transient double vision/absence of ocular motility, transient reduced acuity, transient field defect or transient diplopia. Furthermore, non-cranial ischaemic manifestations were defined as: extra-cranial complications such as fixed vascular stenosis to limb at presentation (secondary to GCA); and extra-cranial ischaemic features such as leg claudication or arm claudication.

Details of the quality control (QC) of GCA case genetic data used for PRS optimization in this work has previously been described in Carmona et al. (2017). Briefly, deoxyribonucleic acid (DNA) was sequenced for UK GCA Consortium samples in three batches. For the first two batches (batch 1 *N*=477, batch 2 *N* =239, of 1,858 total) of genomic DNA were extracted from peripheral blood cells of subjects and genotyped using the Illumina “Infinium HumanCore Beadchip” and the Genotyping Module (v.1.9) of the GenomeStudio software (Illumina).^7^ For these two batches, plus the first batch of WTCCC samples (1958 British birth cohort), several variant QC filters were applied to the data in PLINK v1.07.,^16^ removing single nucleotide polymorphisms (SNPs) with call rates < 0.98, minor allele frequency (MAF) < 0.01, and variants that deviated from Hardy-Weinberg equilibrium (HWE) at *P* < 10^−10^ (for cases) or P < 10^-6^ (for controls). Sex chromosomes were also removed from analyses. Sample QC removed individuals with a sample missingness rate > 5%, and one of each pair of first-degree relatives (identity by descent [IBD] > 0.4) was removed. Principal component analysis (PCA) was performed in PLINK v.1.09.^16^ to account for population stratification in subsequent analyses. Imputation was performed using Minimac4 through the Michigan imputation server^39^. Following imputation, further variant filters were applied, including removal of SNPs with MAF < 0.01 and with imputation quality *r^2^* < 0.5.

An additional 1,142 samples (batch 3) were later recruited to the UK GCA Consortium and genotyped using the Illumina Infidium “Global Screening Array”. For this batch, plus the second WTCCC sample cohort (UK blood service control group), sex chromosomes were removed from analyses and several variant QC filters were applied to the data in PLINK v1.09, removing SNPs with call rates < 0.98, MAF < 0.002, and variants that deviated from the Hardy-Weinberg equilibrium (HWE) at *P* < 10^−10^ (for cases) or P < 10^-6^ (for controls). Sample QC removed individuals with a sample missingness rate > 3%, those who deviated > 3 standard deviations from the heterozygosity rate mean, and all samples with pi-hat (*^p^^*) > 0.2 were removed following linkage disequilibrium (LD) based pruning (high inversion rate regions removed, *window size* [*bp*] = 50, *window shift* [*SNPs*] = 5, *LD R^2^* = 0.2). Principal component analysis (PCA) was performed in PLINK v.1.09. to account for population stratification in subsequent analyses.Imputation was performed using TopMed.^40^

#### Quality control of UK Biobank genetic data

A comprehensive description of the QC of UK Biobank (UKB) genetic data used in this study may be found in Crossfield et al. (2022)^41^ and Bycroft et al. (2018)^19^. Briefly, participants were genotyped using either the Affymetrix UK BiLEVE Axiom or Affymetrix UK Biobank Axiom array and imputation was performed using combined reference panels from the Haplotype Reference Consortium^42^, 1000 Genomes^43^, and UK10K projects.^44^ Sample filters included the removal of individuals above the heterozygosity rate mean of 0.19, high missingness rates (>5%), mismatching genetic/reported sex, and one sample from each pair of those estimated to be second-degree relatives or closer (preferentially removing that with the greater genotype missingness rate). Post imputation QC filters included the removal of samples with *MAF* < 0.001 and imputation quality *r^2^* < 0.8.

Following PCA, the “aberrant” routine in R was employed to detect ethnic outliers (non-Europeans) in PCs 1 and 2, using a lambda parameter of 100. Participants who i) fell within the European PC cluster and ii) self-reported “White” in UKB baseline data (field 21000) were retained for analysis, increasing the “white European” cohort size compared with UKB’s definition, which excludes samples who self-report as “Irish” or “any other white background”.

#### Protein selection

Proteins were selected for study in this work from publicly available summary statistics generated from GWAS investigating inter-individual variation of protein levels.^13^ These analyses were performed using protein quantification data from the multiplexed SOMAscan platform,^45^ consisting of 3,283 modified aptamers (single stranded DNA SOMAmer reagents) for 2,994 proteins/protein complexes (post QC). Details of population demographics, genotyping, protein profiling, QC and statistical analyses of these GWAS have previously been described.^13^ Briefly, informed consent was provided by healthy participants of the INTERVAL study,^46^ which consisted of two non-overlapping sub-cohorts of blood donors aged ≥18 (*N*= 2,731 and 831). The Affymetrix Axiom UK Biobank genotyping array was used to genotype participants and imputation was performed using the Sanger imputation server, using a combined 1000 Genomes Phase 3-UK10K reference panel.

In order to prioritise the proteins investigated and minimise multiple testing, two filters were applied to the selection of plasma proteins for inclusion in the study. Firstly, 730 proteins deemed part of the “human blood secretome” were selected, based on prior knowledge that circulating proteins represent useful druggable targets and because their primary physiological action is in circulating form^15^. Secondly, proteins with low levels of heritability, and therefore unlikely to demonstrate strong genetic associations with GCA-risk, were removed from the study. Heritability (R^2^) estimates (in the form of proportion of variance explained by genome-wide significant variants) were yielded from Sun et al. (2017),^13^ and only those proteins with heritability values greater than or equal to the lowest quartile R^2^ estimate for the group were included in further analyses. After the removal of variants with a low heritability level, 169 proteins remained for analysis.

#### Polygenic risk scoring

The PRSice v2.3.3^17^ software was used for optimizing PRS and testing for associations with GCA. PRS were constructed via the “clumping and thresholding” approach, using effect sizes and *P-values* from GWAS of protein levels.^13^ SNPs were thinned in 500kb blocks, based on an LD R^2^ threshold of 0.1. All SNPs beneath a specified *P-value* threshold (P_T_) in the protein summary statistics were used to form a risk score, which was regressed on the GCA dataset, using the top 10 PCs from PCA as covariates. This process was repeated for risk scores at many P_T_ (minimum P_T_ = 5 x 10^-8^; step size = 5 x 10^-5^; maximum P_T_ = 1). The “best fit” risk score was defined as the PRS with the greatest GCA model fit (*R^2^*) in logistic regression.

A permutation test was used to account for multiple testing within each analysis. This was conducted with PRSice v2.3.3^17^ using the following steps: (1) a “best-fit” risk score was generated; (2) the phenotype was randomly shuffled, the analysis was repeated and another “best-fit” score was generated; (3) step 2 was repeated 10,000 times; and (4) the empirical *p-value* was calculated as

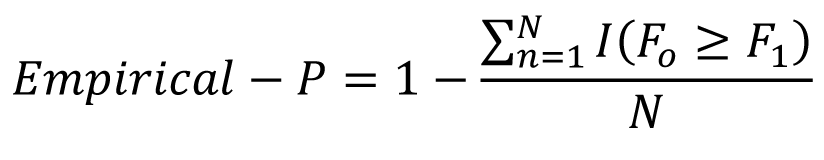

where *I* (.) is the indicator function, *P_o_* is the best-fit P_T_ and *N* is the number of times the analysis was repeated.

To account for multiple testing of numerous proteins, the significance threshold was adjusted using Bonferroni correction. For the 169 proteins assessed by PRS analysis, the corrected *P-value* threshold was 0.05/169 = 2.96 x 10^-4^.

For proteins with a statistically significant PRS, risk scoring was repeated in high-resolution (using a smaller step size between each P_T_ tested; P_T_ step size=5 x 10^-8^ as opposed to 5 x 10^-5^), to increase the precision of the PRS. Variants of the risk score were classed as *cis*-pQTL if they were located within 1MB of the transcription start site of the gene encoding the protein; whilst variants located outside of this region were classed as *trans*-pQTL.

#### Polygenic Risk Score Analysis Using Different Case Definitions

To determine whether there was a difference in the average PRS between those with a “confirmed” GCA diagnosis (via imaging or biopsy) or fulfilling American College of Rheumatology [ACR] classification criteria (with an ACR score >3) and those without, a t-test was performed to assess whether there was a significant difference between the mean PRS at the best fit P_T_ (P_T_ = 1.66 x 10^-3^) in a case-control approach (**Supplementary Table 1**). All individuals in this cohort had a clinical diagnosis of GCA.

To reduce bias from differences in the sample sizes of the groups, a permutation test was applied. A t-test was performed on the target phenotype to retrieve a test statistic. Phenotype values were then randomly re-assigned and a t-test performed again. This was repeated 10,000 times, creating an empirical distribution of the test statistic under the null hypothesis. Finally, an empirical *P-value* was calculated as:

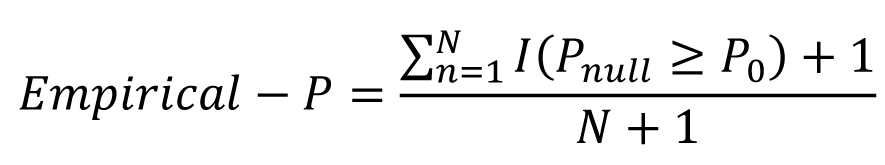

Where *I(.)* is the indicator function, *F_0_*is the *F-value* using the random sample, *F_1_* is the *F-value* using the initial sub-sample and *N* is the number of times the analysis was repeated.

#### Testing PRS associations with giant cell arteritis severity

To determine whether PRS associated with GCA susceptibility were also predictive of GCA severity, PRS which had a statistically significant association with GCA at the Bonferroni-corrected threshold (*P-value*=2.96 x 10^-4^) were tested for association with a proxy for GCA severity: cranial ischaemic complications in GCA. This was performed in the GCA cohort with detailed clinical data available, with cases defined as GCA patients with reported cranial ischaemic complications and controls defined as GCA patients without reported cranial ischaemic complications (cases *N*=317, controls *N*=1,542). Two other proxies for disease severity were also used: transient cranial ischaemic manifestations in GCA (cases *N*=1,127, controls *N*=719), and non-cranial ischaemic features in GCA (cases *N*=195, controls *N*=1,371).

#### Mendelian randomization

To investigate the causality of associated proteins in GCA, case-control summary statistics (in the form of log odds ratios) were combined with summary statistics for associated proteins in MR.

Three genetic scores were constructed for use in MR, each with low levels of LD (R^2^ < 0.1), and with RS numbers taken from Ensembl’s Variant Effect Predictor (VEP)^47^. The inverse-variance weighted (IVW) method was used to perform two-sample MR, and this was repeated using the weighted median and weighted mode methods. Sensitivity analyses (including Cochran’s heterogeneity test and the MR-Egger intercept test) were performed on the scores to rule out potential horizontal pleiotropy, which could invalidate use of the genetic instruments^14^.

Potentially pleiotropic variants in MR sensitivity analyses were further investigated using a PheWAS approach. Genome-wide associations (defined as associations with a *p-value* ≤ 5 x 10^-8^) were collated from three databases: GWAS Catalog^48^, Phenoscanner^49^, and the MR-Base PheWAS catalogue of summary data^50^. Traits associated with ≥2 SNPs were used as the exposure variable in MR (using individual SNPs as genetic instruments with the Wald ratio method), to test for a causal association between these traits and GCA via the SNPs in question. MR and sensitivity analyses were then repeated with outliers removed.

## Supplementary Results

### Polygenic risk scoring borderline significant results

A small number of protein PRS had associations with GCA which passed the *P*<0.05 *P-value* threshold but not the Bonferroni-corrected threshold. These PRS were denoted “borderline” proteins in this work.

The interleukin 1 receptor type 2 (IL1R2) PRS had an association with GCA with a positive direction of effect (*coefficient*=15.29; *standard error*[*SE*]=3.97; *P-value*[*P*]=2.6 x 10^-3^). This PRS consisted of 487 SNPs (*P_T_*=3 x 10^-4^) and had a model fit of *R^2^*=0.004. Furthermore, a superoxide dismutase 3 (SOD3) PRS, consisting of 388 SNPs (*P_T_*=2.5 x 10^-4^), had a GCA association with a positive direction of effect (*coefficient*=15.36; SE=4.03; *P*=4.2 x 10^-3^). This PRS had a model fit of *R^2^*=0.004. The best-fit C-C motif chemokine ligand 22 (CCL22) PRS had an association with GCA with a positive direction of effect (*coefficient*=10.04; *SE*=2.89; *P*=0.01). This PRS had a model fit of *R^2^*=0.004 and consisted of 254 SNPs (*P_T_*=1.5 x 10^-4^).

Additionally, a coagulation factor X (F10) PRS, comprising 114,475 SNPs (*P_T_*=0.19) had an association with GCA with a positive direction of effect (*coefficient*=366.13; *SE*=106.2; *P*=0.01). This PRS had a model fit of *R^2^*=0.003.

The serpin family G member 1 (SERPING1, otherwise known as C1-inhibitor) PRS had an association with GCA with a positive direction of effect (*coefficient*=382.35; *SE*=112.25; *P*=0.01). This PRS consisted of 130,831 SNPs (*P_T_*=0.23) and had a model fit of R^2^=0.003. The 11 SNP (*P_T_*=5 x 10^-8^) plasminogen (PLG) PRS was associated with GCA, with a positive direction of effect (coefficient=0.83; *SE*=0.25; *P*=0.01) and a model fit of *R^2^*=0.003. Likewise, the gastrin-releasing peptide (GRP) PRS was also associated with GCA (*coefficient*=-0.49; *SE*=0.15; *P*=0.02). The 2 SNP PRS (*P_T_*=5 x 10^-8^) had a model fit of *R^2^*=0.003 and a negative direction of effect.

The C1Q and TNF related 1 (C1QTNF1) PRS was associated with GCA, with a positive direction of effect (*coefficient*=294.32; *SE*=90.47; *P*=0.02). This 91,757 SNP PRS (*P_T_*=0.14) had a model fit of *R^2^*=0.003. The vitronectin (VTN) PRS, comprised of 9,792 SNPs (*P_T_*=8.75 x 10^-3^), had an association with GCA with a negative direction of effect (*coefficient*=-72; *SE*=22.46; *P*=0.02). This PRS had a model fit of *R^2^*=0.003. Likewise, the 2,099 SNP (*P_T_*=1.55 x 10^-3^) TNF superfamily member 12 (TNFSF12) PRS had a GCA association with a negative direction of effect (*coefficient*=-30.48; *SE*=10.1; *P*=0.04), with a model fit of *R^2^*=0.003. Finally, the melanoma inhibitory activity (MIA) PRS, consisting of 16,226 SNPs, had an association with GCA with a positive direction of effect (*coefficient*=90.59; *SE*=31.03; *P*=0.05). This PRS had a model fit of *R^2^*=0.003.

### High-resolution APOL1 scoring

To increase the precision of the APOL1 risk score, risk scoring was performed at greater resolution, by reducing the step size between *P-value* thresholds (**Supplementary Figure 1**). Results revealed that the best-fit P_T_ was marginally smaller than that found in the first analysis and consisted of 51 fewer SNPs (*P_T_*=1.66 x 10^-3^; *N*=2,227). Additionally, the regression coefficient of the relationship was more extreme (*coefficient*=-320.49, *SE*=60.22).

### Polygenic Risk Score Analysis Using Different Case Definitions

To assess whether individuals with greater diagnostic confidence demonstrate a stronger genetic propensity to GCA through the APOL1 PRS, the case cohort was stratified based on the presence of a confirmed diagnosis, and the average PRS at the optimal P_T_ was compared.

No statistically significant difference in the APOL1 PRS was observed between those with a positive biopsy and/or imaging result, and those without a positive biopsy or imaging result (t[785.55]=-0.23, *P*=0.41, positive biopsy/imaging mean = -0.08, negative biopsy/imaging mean= -0.09). A similar observation was made when comparing those with an ACR score ≥ 3 and those with an ACR score < 3 (t[102.16]=0.1, *P*=0.54, positive biopsy/imaging mean=-0.06; negative biopsy/imaging mean=-0.08).

### Testing PRS associations with secondary outcomes

To determine whether those PRS associated with GCA susceptibility were also predictive of GCA severity, PRS were tested for associations with several ischaemic phenotypes in GCA, proxies for disease severity (**Supplementary Table 2**). The APOL1 PRS did not demonstrate a statistically significant association with cranial ischaemic complications (*odds ratio*[*OR*]=0.76, 95% *confidence intervals*[*CIs*]=0.51 to 1.12, *P*=0.324, for the highest versus lowest APOL1 quintile), transient cranial ischaemic manifestations (*OR*[95% *CIs*]=1.15 [0.85 to 1.55], *P*=0.53, for the highest versus lowest APOL1 quintile), or non-cranial ischaemic features (*OR*[95% *CIs*]=1.15 [0.85 to 1.55], *P*=0.53, for the highest versus lowest APOL1 quintile).

### Mendelian randomization

In MR, genetic variants are used as instrumental variables (IVs) to assess the causal relationship between an exposure (e.g. protein) and an outcome (e,g. GCA-risk). Three genetic scores were constructed for use as IVs in this study: a “liberal” score (APOL1 *P-value* ≤ 1 x 10^-4^), an “intermediate” score (APOL1 *P-value* ≤ 5 x 10^-5^), and a “conservative” score (APOL1 *P-value* ≤ 5 x 10^-8^). A small number (n = 2) of variants were lost from the scores due to the inability of VEP to generate SNP RS numbers.

Once effect and non-effect alleles were harmonised between the exposure (pQTL) and outcome (GCA-risk) data, MR analysis was performed using each of the three risk scores. Using the IVW method of MR, the APOL1 protein was estimated by all risk scores to have a statistically significant, causal effect on GCA-risk (**Supplementary Table 3**). This direction of effect was always negative (liberal *beta*[*SE*]=-0.093[0.02]; intermediate *beta*[*SE*]=-0.131[0.05]; conservative *beta*[*SE*]=-0.22[0.1]), and was supported by the weighted median and weighted mode methods of MR (**Supplementary Figure 3**); although statistical significance was only found using the weighted median method with the liberal score, and the weighted mode method with the conservative score.

Results of the MR-Egger intercept test lacked statistical significance for any of the risk scores (**Supplementary Table 4**), indicating that directional horizontal pleiotropy of SNPs in the risk score is unlikely to have affected the effect size or direction of estimates. However, there was evidence for pleiotropy in the conservative score (Cochran’s *Q*= 24.8; *degrees of freedom*=10; *P- value*=5.71 x 10^-3^).

SNPs from the conservative score (*N*=11) were investigated using a PheWAS in order to identify associations with other proteins that could be acting as alternate causal pathways to GCA. PheWAS results revealed 19 proteins/traits that have a statistically significant association with two or more variants (**Supplementary Table 5**). These traits were used as exposure variables to assess their causal association with GCA in MR. 14 results (across 3 SNPs: rs11599750, rs5167 and rs704) revealed a statistically significant causal effect of the trait on GCA-risk, in the same direction as the variant’s direct association with GCA-risk (**Supplementary Table 6**). These SNPs are located on different chromosomes (chromosomes 10, 19 and 17, respectively), so LD is unlikely to be affecting these associations. Such results provide evidence that these variants could be affecting GCA-risk through a pathway other than APOL1, and may therefore be inappropriate genetic instruments for use in APOL1 MR. It should be noted that when multiple testing correction is applied to these analyses using the Bonferroni method, none of the associations identified would have passed the 0.05/53 = 9.43 x 10^-4^ *P-value* threshold.

Once the three outliers (rs704, rs11599750 and rs5167) were removed, MR was repeated with each risk score. Using the IVW method of MR, the causal effect of the APOL1 protein on GCA risk remained statistically significant for each of the risk scores, with a negative direction of effect. Sensitivity analyses revealed that horizontal pleiotropy was now unlikely to be affecting the causal estimate (**Supplementary Table 7**).

## UK GCA Consortium

**Management Team:** Ann W Morgan, Sarah L Mackie, Louise Sorensen **Laboratory Team:** Lubna Haroon Raashid, Steve Martin, James I Robinson School of Medicine, University of Leeds, Leeds UK

## Recruiting Sites

Chapel Allerton Hospital, Chapeltown Road, Leeds, LS7 4SA, UK PI: Ann Morgan, Sarah Mackie

Research staff: Oliver Wordsworth, Isobel Whitwell, Jessica Brock

Pinderfields General Hospital, Aberford Road, *Wakefield*, WF1 4DG, UK PI: Victoria Douglas, Chamila Hettiarachchi

Research staff: Jacqui Bartholomew

Dewsbury District and General Hospital, Halifax Road, Dewsbury, WF13 4HS, UK PIs: Stephen Jarrett, Gayle Smithson, Chamila Hettiarachchi

Research staff: Jacqui Bartholomew

York Hospital, Wigginton Road, York, YO31 8HE, UK PI: Michael Green

Research staff: Pearl Clark Brown

Harrogate District Hospital, Lancaster Park Rd, Harrogate, HG2 7SX, UK PI: Cathy Lawson

Research staff: Esther Gordon

Ipswich Hospital, Heath Road, Ipswich, IP4 5PD, UK PI: Suzanne Lane

Research staff: Rebecca Francis

Southend Hospital, Prittlewell Chase, Westcliff-on-Sea, Essex, SS0 0RY, UK PI: Bhaskar Dasgupta, Bridgett Masunda

Research staff: Jo Calver

Hull Royal Infirmary, Anlaby Road, *Hull*, HU3 2JZ, UK PI: Yusuf Patel

Research staff: Charlotte Thompson, Louise Gregory

Croydon University Hospital, 530 London Road, Croydon, Surrey, CR7 7YE, UK PI: Sarah Levy

Haywood Hospital, High Lane, Burslem, Staffordshire, ST6 7AG, UK PI: Ajit Menon

Research staff: Amy Thompson, Lisa Dyche, Michael Martin

Royal Surrey County Hospital, Egerton Road, Guildford, GU2 7XX, UK PI: Charles Li

Queen’s Hospital, Belvedere Road, Burton-on-Trent, DE13 0RB, UK PI: Ramasharan Laxminarayan

Research staff: Louise Wilcox, Ralph de Guzman

Freeman Hospital, Freeman Road, High Heaton, Newcastle upon Tyne, Tyne and Wear, NE7 7DN, UK

PI: John Isaacs, Alice Lorenzi, Gary Reynolds Research staff: Ross Farley, Helain Hinchcliffe-Hume

Barnsley Hospital, Gawber Road, Barnsley, S75 2EP, UK PI: Victoria Bejarano

Research staff: Susan Hope

Northampton General Hospital, Cliftonville, Northampton, NN1 5BD, UK PI: Pradip Nandi

Research staff: Lynne Stockham, Catherine Wilde, Donna Durrant

Frimley Park Hospital, Portsmouth Road, Camberley, GU16 7UJ, UK PI: Mark Lloyd

Doncaster Royal Infirmary, Armthorpe Road, Doncaster, DN2 5LT, UK PI: Chee-Seng Ye, Rob Stevens

Chelsea and Westminster Hospital, Fulham Road, London, SW10 9NH, UK PI: Amjad Jilani

Great Western Hospital, Marlborough Road, Swindon, SN3 6BB, UK PI: David Collins

Research staff: Suzannah Pegler, Ali Rivett, Liz Price

Royal National Hospital for Rheumatic Diseases, Upper Borough Walls, Bath, Avon, BA1 1RL, UK

PI: Neil McHugh, Sarah Skeoch

Research staff: Diana O’Kane, Sue Kirkwood

Gateshead Queen Elizabeth Hospital, Queen Elizabeth Avenue, Gateshead, NE9 6SX, UK PI: Saravanan Vadivelu

Research staff: Susan Pugmire

Airedale General Hospital, Skipton Road, Keighley, BD20 6TB, UK PI: Shabina Sultan

Research staff: Emma Dooks, Lisa Armstrong

Warrington Hospital, Lovely Lane, Warrington, Cheshire, WA5 1QG, UK PI: Hala Sadik

Basildon University Hospital, Nethermayne, Basildon, SS16 5NL, UK PI: Anupama Nandagudi

Research staff: Tolu Abioye, Angelo Ramos, Steph Gumus

St George’s Hospital, Blackshaw Road, London, SW17 0QT, UK PI: Nidhi Sofat

Research staff: Abiola Harrison, Abi Seward

Queen Elizabeth Hospital, Mindelsohn Way, Birmingham, B15 2TH, UK PI: Susan Mollan

Research staff: Ray Rahan, Helen Hawkins

Royal Preston Hospital, Sharoe Green Lane, Preston, PR2 9HT, UK PI: Hedley Emsley, Anna Bhargava

Research staff: Vicki Fleming, Marianne Hare, Sonia Raj

Arrowe Park Hospital, Arrowe Park Road, Birkenhead, CH49 5PE, UK PI: Emmanuel George

Research staff: Nicola Allen, Karl Hunter

King’s College Hospital, Denmark Hill, London, SE5 9RS, UK PI: Eoin O’Sullivan

Research staff: Georgina Bird

Stoke Mandeville Hospital, Mandeville Road, Aylesbury, HP21 8AL, UK PI: Malgorzata Magliano

Research staff: Katarina Manzo, Bobbie Sanghera

Royal Cornwall Hospital, Treliske, Truro, TR1 3LJ, UK PI: David Hutchinson

Research staff: Fiona Hammonds

Peterborough City Hospital, Bretton Gate, Peterborough, PE3 9GZ, UK PI: Poonam Sharma

Research staff: Richard Cooper, Graeme McLintock

Scarborough Hospital, Woodlands Drive, Scarborough, YO12 6QL, UK PI: Zaid S. Al-Saffar, Mike Green

Research staff: Kerry Elliott, Tania Neale, Janine Mallinson Queen’s Medical Centre, Derby Road, Nottingham, NG7 2UH, UK

PI: Peter Lanyon

Research staff: Marie-Josephe Pradere

Addenbrooke’s Hospital, Hills Road, Cambridge, CB2 0QQ, UK PI: Natasha Jordan, Ei Phyu Htut

Research staff: Thelma Mushapaidzi, Donna Abercrombie, Sam Wright, Jane Rowlands

Norfolk and Norwich University Hospital, Colney Lane, Norwich, NR4 7UY, UK PI: Chetan Mukhtyar

Research staff: James Kennedy

James Paget Hospital, Lowestoft Road, Great Yarmouth, NR31 6LA, UK PI: Damodar Makkuni

Research staff: Elva Wilhelmsen

The Queen Elizabeth Hospital, Gayton Road, King’s Lynn, PE30 4ET, UK PI: Michael Kouroupis

West Suffolk Hospital, Hardwick Lane, Bury St Edmunds, IP33 2QZ, UK PI: Shweta Bhagat

Research staff: Lily John

Ashford and St. Peter’s Hospitals, London Road, Ashford, TW15 3AA, UK PI: Rod Hughes

Research staff: Margaret Walsh, Marie Buckley

Torbay Hospital, Newton Road, Torquay, 7AA, UK PI: Kirsten Mackay

Research staff: Tracey Camden-Woodley, Joan Redome, Kirsty Pearce

Lister Hospital, Coreys Mill Lane, Stevenage, SG1 4AB, UK PI: Thiraupathy Marianayagam

Research staff: Carina Cruz, Elizabeth Warner

North Tyneside General Hospital, Rake Lane, North Shields, NE29 8NH, UK PI: Ishmael Atchia

Research staff: Claire Walker, Karen Black, Stacey Duffy

Royal Lancaster Infirmary/Westmorland General Hospital, Ashton Road, Lancaster, LA1 4RP, UK

PI: Marwan Bukhari

Research staff: Lynda Fothergill, Rebecca Jefferey, Jackie Toomey

Royal Glamorgan Hospital, Ynysmaerdy, Pontyclun, CF72 8XR, UK PI: Ceril Rhys Dillon

Research staff: Carla Pothecary, Lauren Green

Warwick Hospital, Lakin Road, Warwick, CV34 5BW, UK PI: Tracey Toms

Research staff: Linda Maher

West Middlesex Hospital, Twickenham Road, Isleworth, TW7 6AF, UK PI: Diana Davis

Research staff: Amrinder Sayan, Mini Thankachen

Royal Devon and Exeter Hospital, Barrack Road, Exeter, EX2 5DW, UK PI: Mahdi Abusalameh

Research staff: Jessica Record

Birmingham Heartlands Hospital, Bordesley Green, Birmingham, B9 5SS, UK PI: Asad Khan

Research staff: Sam Stafford

Royal Derby Hospital, Uttoxeter Road, Derby, DE22 3NE, UK PI: Azza Hussein

Research staff: Clare Williams, Alison Fletcher, Laura Johson, Richard Burnett

Aintree University Hospital, Lower Lane, Liverpool, L9 7AL, UK PI: Robert Moots

Research staff: Helen Frankland

University Hospital Wishaw, Netherton Street, Wishaw, ML2 0DP, UK PI: James Dale

Research staff: Karen Black, Kirsten Moar, Carol Hollas

Manchester Royal Infirmary, Oxford, Manchester, M13 9WL, UK PI: Ben Parker

Research staff: Derek Ridings, Sandhya Eapen, Sindhu John

Bristol Royal Infirmary, Upper Maudlin Street, Bristol, BS2 8HW, UK PI: Jo Robson

Research staff: Lucy Belle Guthrie, Rose Fyfe, Moira Tait

Christchurch Hospital, Fairmile Road, Christchurch, BH23 2JX, UK PI: Jonathan Marks

Research staff: Emma Gunter, Rochelle Hernandez

Ninewells Hospital & Medical School, James Arrott Drive, Dundee, DD2 1SG, UK PI: Smita Bhat

Research staff: Paul Johnston

Poole Hospital, Longfleet Road, Poole, BH15 2JB, UK

PI: Muhammad Khurshid Research staff: Charlotte Barclay

Countess of Chester Hospital, Liverpool Road, Chester, CH2 1UL, UK PI: Deepti Kapur

Research staff: Helen Jeffrey, Anna Hughes, Lauren Slack

Nevill Hall Hospital, Brecon Road, Abergavenny, NP7 7EG, UK PI: Eleri Thomas

Research staff: Anna Royon, Angela Hall

Derriford Hospital, Derriford Road, Plymouth, PL6 8DH, UK PI: Jon King

Research staff: Sindi Nyathi

University College Hospital, Euston Road, London, NW1 2BU, UK PI: Vanessa Morris, Madhura Castelino

Research staff: Ellie Hawkins, Linda Tomson

Royal Free Hospital, Pond Street, London, NW3 2QY, UK PI: Animesh Singh

Research staff: Annalyn Nunag, Stella O’Connor

Darlington Memorial Hospital, Hollyhurst Road, Darlington, DL3 6HX, UK PI: Nathan Rushby

Research staff: Nicola Hewitson

Leicester Royal Infirmary, Infirmary Square, Leicester, LE1 WW, UK PI: Kenny O’Sunmboye

Research staff: Adam Lewszuk, Louise Boyles

Royal Alexandra Hospital/Vale of Leven Hospital/Inverclyde Royal Hospital, Great Western Road, Glasgow, G12 0XH, UK

PI: Martin Perry

Royal Hampshire County Hospital, Romsey Road, Winchester, SO22 5DG, UK PI: Emma Williams

Research staff: Christine Graver, Emmanuel Defever

Salford Royal Hospital, Stott Lane, Salford, M6 8HD, UK PI: Sanjeet Kamanth

Research staff: Dominic Kay, Joe Ogor, Louise Winter

Minerva Health Centre, Lowthorpe Rd, Preston PR1 6SB, UK PI: Sarah Horton

Research staff: Gillian Welch, Kath Hollinshead

Hammersmith Hospital, Du Cane Road, London, W12 0HS, UK PI: James Peters

Research nurses: Julius Labao, Andrea Dmello

St Helens Hospital, Marshalls Cross Rd, Saint Helens WA9 3DA, UK PI: Julie Dawson

Research staff: Denise Graham

Queen Elizabeth the Queen Mother Hospital, Ramsgate Rd, Margate CT9 4AN, UK PI: Denise De Lord

Research staff: Jo Deery, Tracy Hazelton

---

**Supplementary Figure 1.**
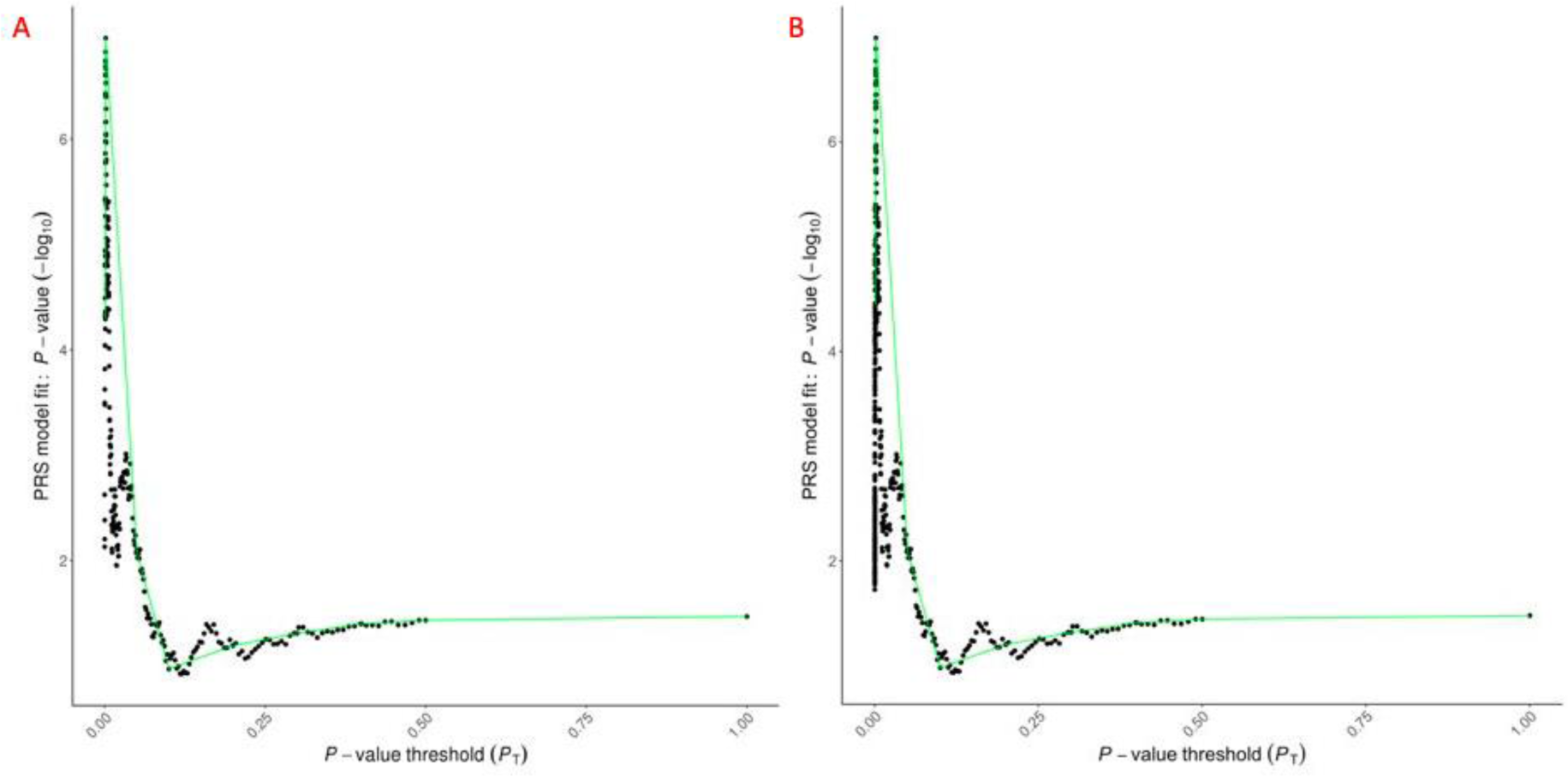
Comparison of high-resolution plots generated in the regression of APOL1 risk scores on GCA-risk. Each plot shows significance values of the model fit (– log_10_ *P-values*) between all polygenic risk scores (PRS) generated from Apolipoprotein L1 (APOL1) protein quantitative trait loci (pQTL) and risk of giant cell arteritis (GCA). The green line connects points demonstrating broad *P-value* thresholds (P_T_) displayed in the corresponding bar plots (see Figure 4.2). A) high-resolution plot generated using a P_T_ step size of 5 x 10^-5^; B) high-resolution plot generated using a P_T_ step size of 5 x 10^-8^.

**Supplementary Figure 3.**
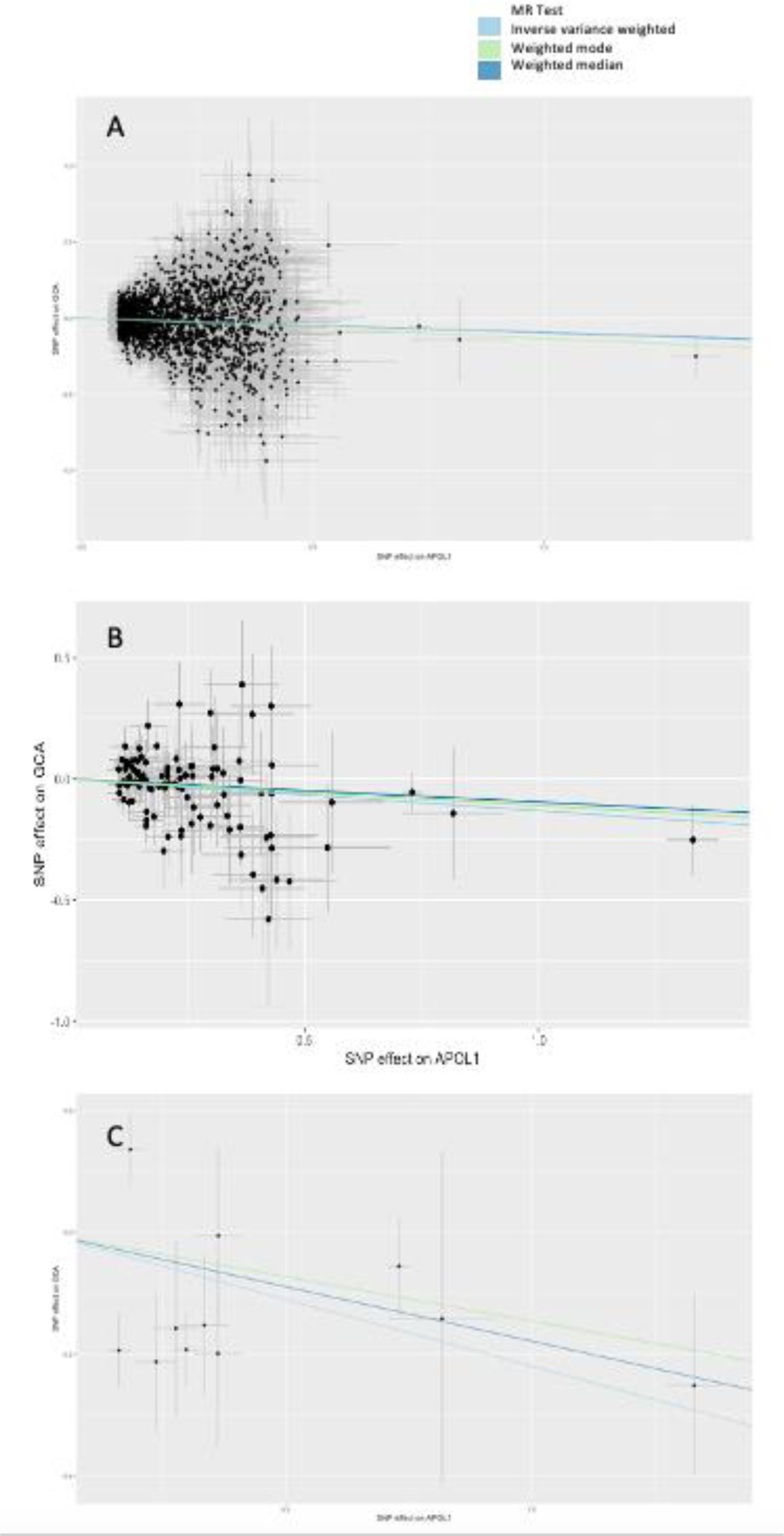
Scatter plot of the per-SNP effect of the (top) liberal (1 x10^-4^), (middle) intermediate (5 x 10^-5^), and (bottom) conservative (*P* < 1 x 10^-4^) Apolipoprotein L1 (APOL1) risk scores on giant cell arteritis (GCA) risk, with the causal estimates of the entire risk score regressed onto the plot and colour coded by method (inverse variance weighted, weighted median and weighted mode MR).

**Supplementary Table 1.** Features of groups compared in PRS sensitivity analyses.

|  | Cases *N* | Controls *N* |
| --- | --- | --- |
| **Biopsy and/or imaging positive vs. Biopsy and imaging negative\*** | 963 | 423 |
| **ACR score $\geq 3$ vs. ACR score $< 3$†** | 1,287 | 92 |

*N*, number; ACR, American college of rheumatology.
\*Individuals without diagnostic tests performed were excluded from this analysis.
†Individuals without documented ACR criteria/scores were excluded from this analysis.

**Supplementary Table 2.**
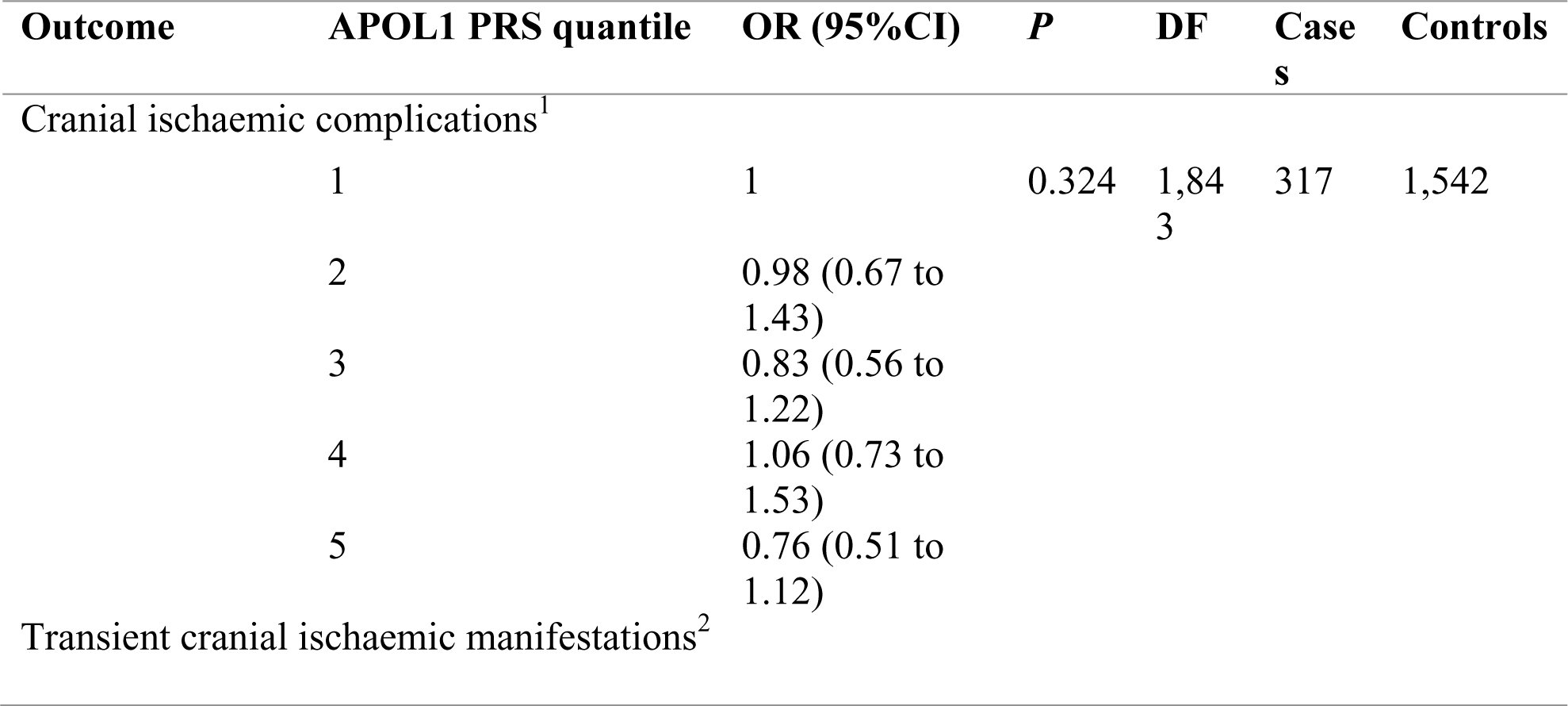
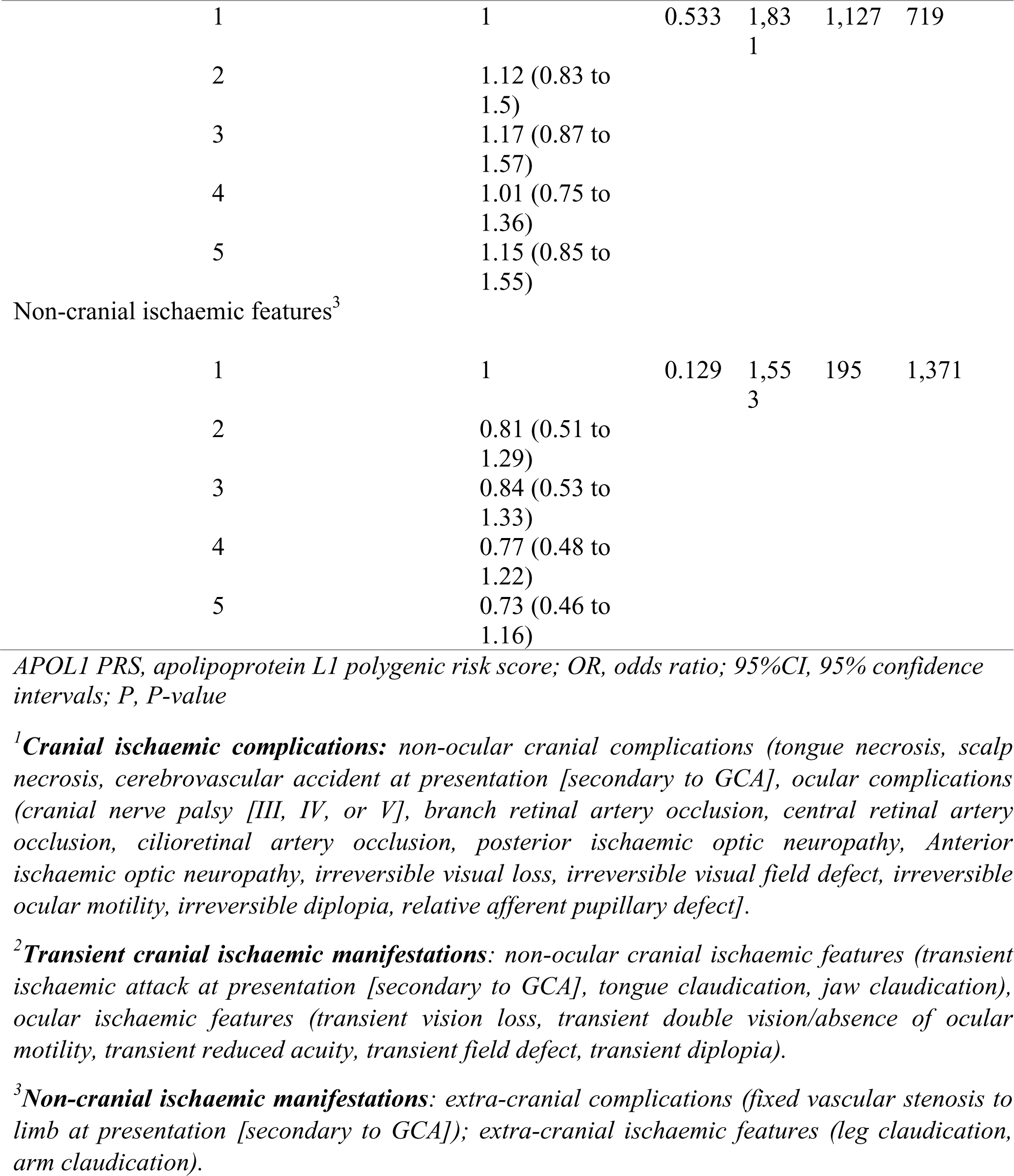
APOL1 polygenic risk score associations with proxies for GCA severity, including: cranial ischaemic complications in GCA, transient cranial ischaemic manifestations in GCA, and non-cranial ischaemic features in GCA.

| Outcome | APOL1 PRS quantile | OR (95%CI) | P | DF | Cases | Controls |
| --- | --- | --- | --- | --- | --- | --- |
| Cranial ischaemic complications<sup>1</sup> |  |  |  |  |  |  |
|  | 1 | 1 | 0.324 | 1,843 | 317 | 1,542 |
|  | 2 | 0.98 (0.67 to 1.43) |  |  |  |  |
|  | 3 | 0.83 (0.56 to 1.22) |  |  |  |  |
|  | 4 | 1.06 (0.73 to 1.53) |  |  |  |  |
|  | 5 | 0.76 (0.51 to 1.12) |  |  |  |  |
| Transient cranial ischaemic manifestations<sup>2</sup> |  |  |  |  |  |  |

|  |  |  |  |  |  |
| --- | --- | --- | --- | --- | --- |
| 1 | 1 | 0.533 | 1,831 | 1,127 | 719 |
| 2 | 1.12 (0.83 to 1.5) |  |  |  |  |
| 3 | 1.17 (0.87 to 1.57) |  |  |  |  |
| 4 | 1.01 (0.75 to 1.36) |  |  |  |  |
| 5 | 1.15 (0.85 to 1.55) |  |  |  |  |
| Non-cranial ischaemic features<sup>3</sup> |  |  |  |  |  |
| 1 | 1 | 0.129 | 1,553 | 195 | 1,371 |
| 2 | 0.81 (0.51 to 1.29) |  |  |  |  |
| 3 | 0.84 (0.53 to 1.33) |  |  |  |  |
| 4 | 0.77 (0.48 to 1.22) |  |  |  |  |
| 5 | 0.73 (0.46 to 1.16) |  |  |  |  |

*APOL1 PRS, apolipoprotein L1 polygenic risk score; OR, odds ratio; 95%CI, 95% confidence intervals; P, P-value*
<sup>1</sup>**Cranial ischaemic complications:** non-ocular cranial complications (tongue necrosis, scalp necrosis, cerebrovascular accident at presentation [secondary to GCA], ocular complications (cranial nerve palsy [III, IV, or V], branch retinal artery occlusion, central retinal artery occlusion, cilioretinal artery occlusion, posterior ischaemic optic neuropathy, Anterior ischaemic optic neuropathy, irreversible visual loss, irreversible visual field defect, irreversible ocular motility, irreversible diplopia, relative afferent pupillary defect).
<sup>2</sup>**Transient cranial ischaemic manifestations:** non-ocular cranial ischaemic features (transient ischaemic attack at presentation [secondary to GCA], tongue claudication, jaw claudication), ocular ischaemic features (transient vision loss, transient double vision/absence of ocular motility, transient reduced acuity, transient field defect, transient diplopia).
<sup>3</sup>**Non-cranial ischaemic manifestations:** extra-cranial complications (fixed vascular stenosis to limb at presentation [secondary to GCA]); extra-cranial ischaemic features (leg claudication, arm claudication).

**Supplementary Table 3.**
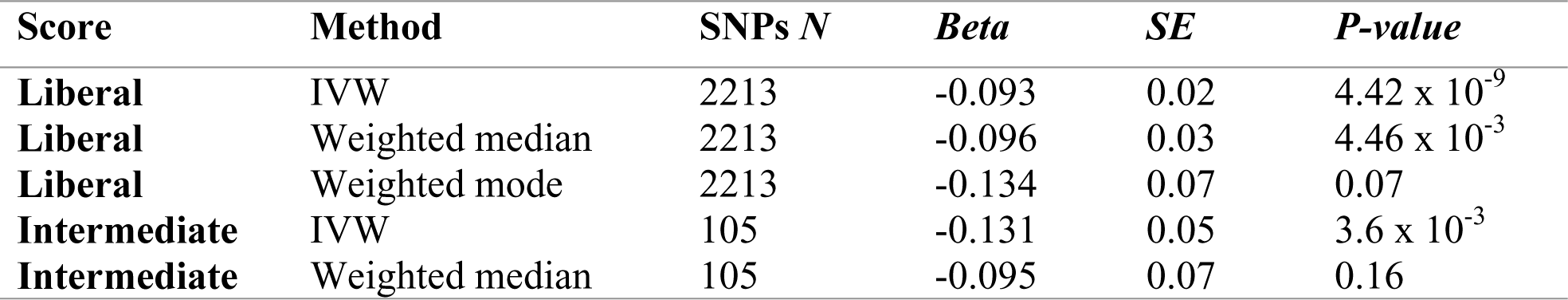
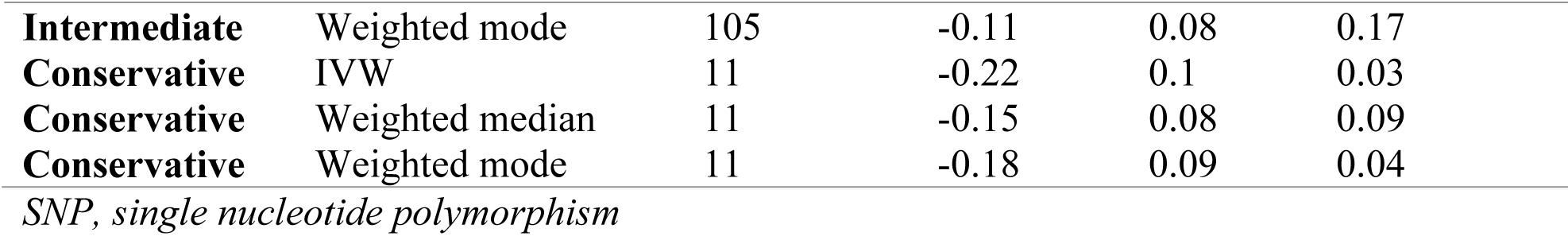
Causal estimation of the effect of Apolipoprotein L1 (APOL1) on giant cell arteritis (GCA) risk using various methods of performing mendelian randomization (MR).

| Score | Method | SNPs *N* | *Beta* | *SE* | *P-value* |
| --- | --- | --- | --- | --- | --- |
| **Liberal** | IVW | 2213 | -0.093 | 0.02 | $4.42 \times 10^{-9}$ |
| **Liberal** | Weighted median | 2213 | -0.096 | 0.03 | $4.46 \times 10^{-3}$ |
| **Liberal** | Weighted mode | 2213 | -0.134 | 0.07 | 0.07 |
| **Intermediate** | IVW | 105 | -0.131 | 0.05 | $3.6 \times 10^{-3}$ |
| **Intermediate** | Weighted median | 105 | -0.095 | 0.07 | 0.16 |
| **Intermediate** | Weighted mode | 105 | -0.11 | 0.08 | 0.17 |
| **Conservative** | IVW | 11 | -0.22 | 0.1 | 0.03 |
| **Conservative** | Weighted median | 11 | -0.15 | 0.08 | 0.09 |
| **Conservative** | Weighted mode | 11 | -0.18 | 0.09 | 0.04 |

*SNP, single nucleotide polymorphism*

**Supplementary Table 4.** Summary of results from MR sensitivity analyses performed using the APOL1 PRS.

| Risk Score | MR-Egger intercept (SE) | MR-Egger intercept $P$-value | Cochran's Q | Cochran's Q DF | Cochran's Q $P$-value |
| --- | --- | --- | --- | --- | --- |
| **Liberal** | -0.005 (0.005) | 0.31 | 2209.1 | 2212 | 0.51 |
| **Intermediate** | 0.03 (0.017) | 0.06 | 101.67 | 104 | 0.55 |
| **Conservative** | -0.07 (0.07) | 0.34 | 24.82 | 10 | $5.71 \times 10^{-3}$ |

**Supplementary Table 5.**
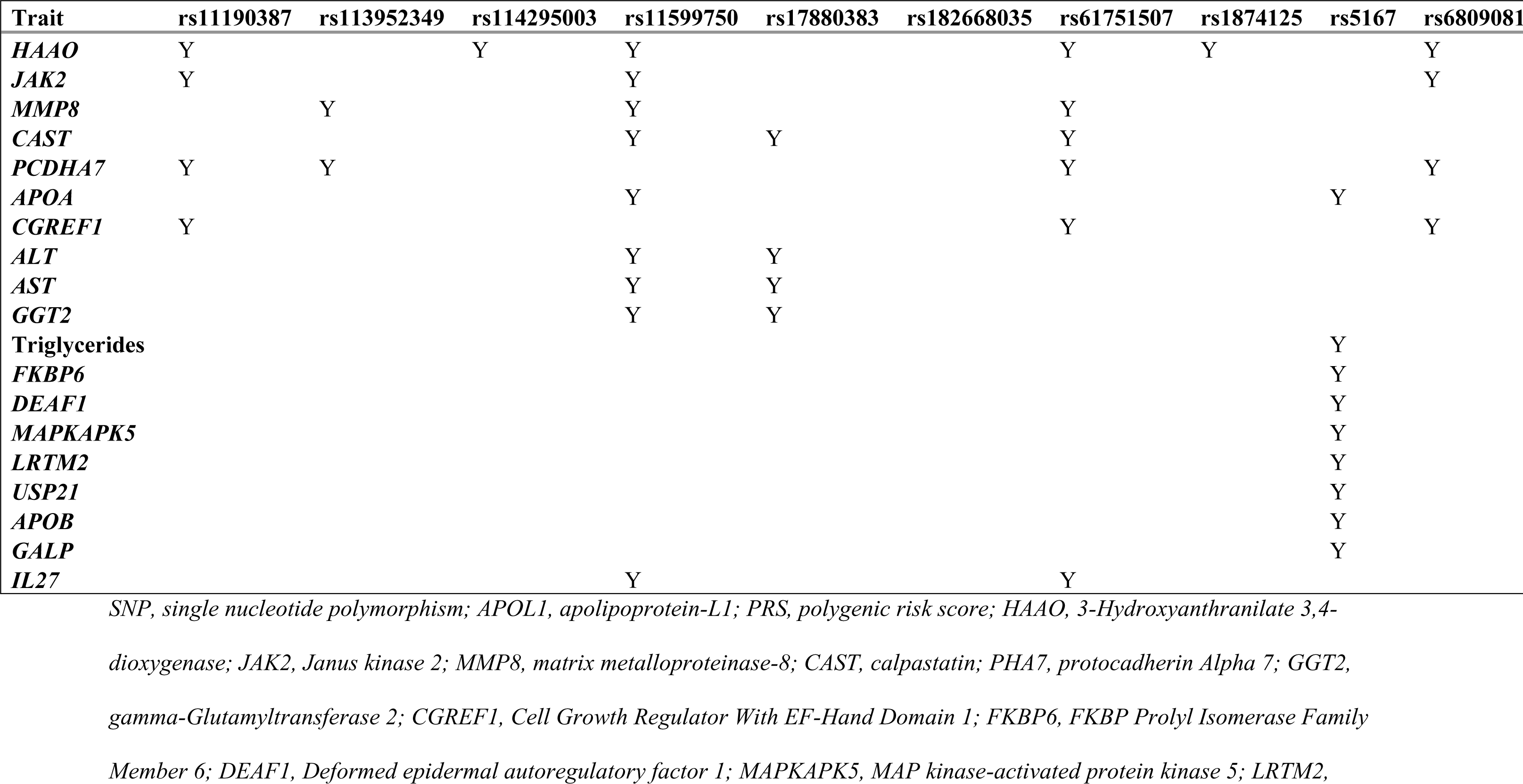
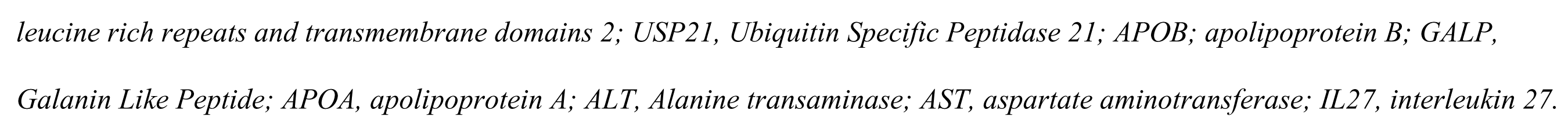
Matrix summarising proteins/traits for which associations with SNPs of the conservative APOL1 PRS were identified.

**Supplementary Table 6.**
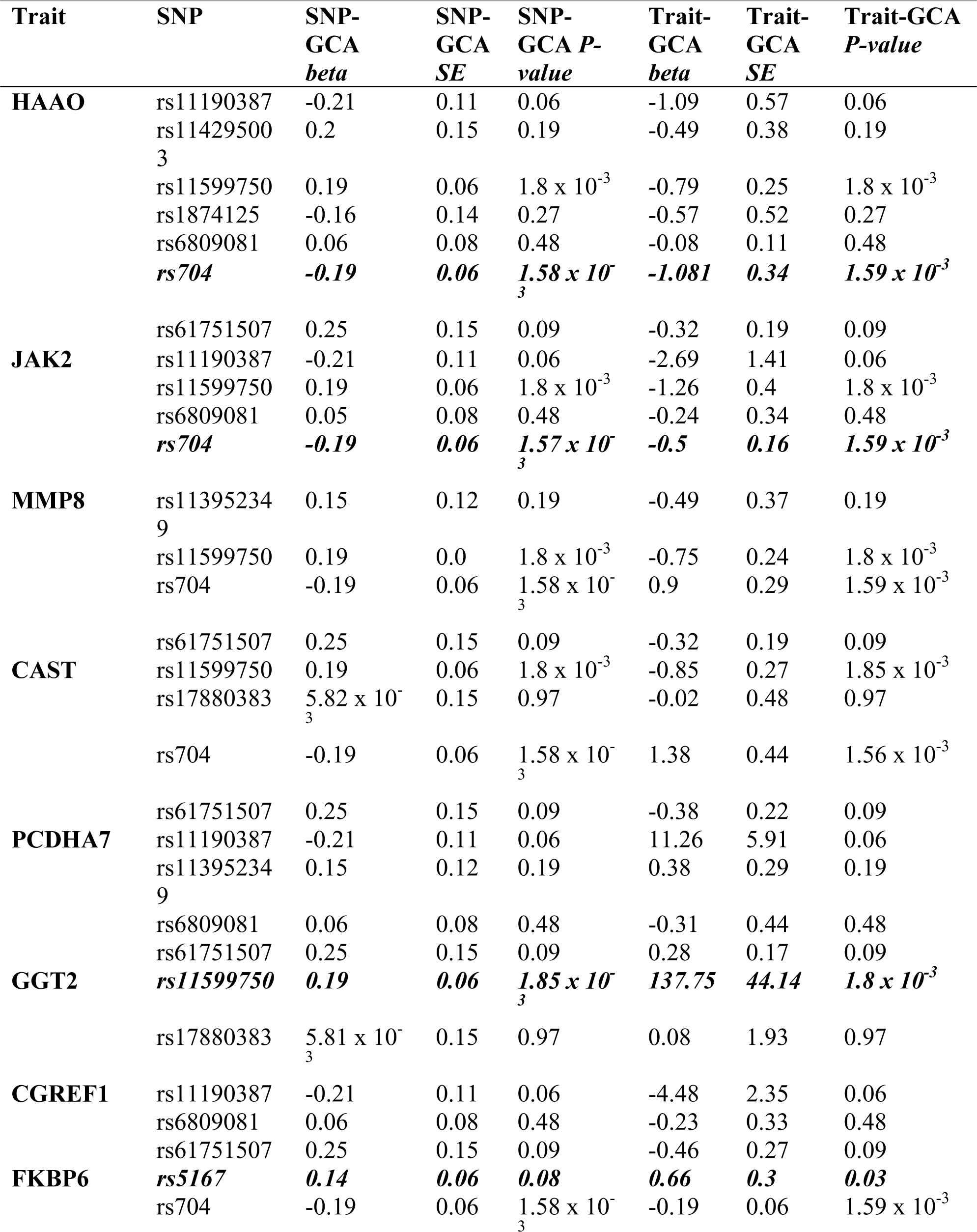
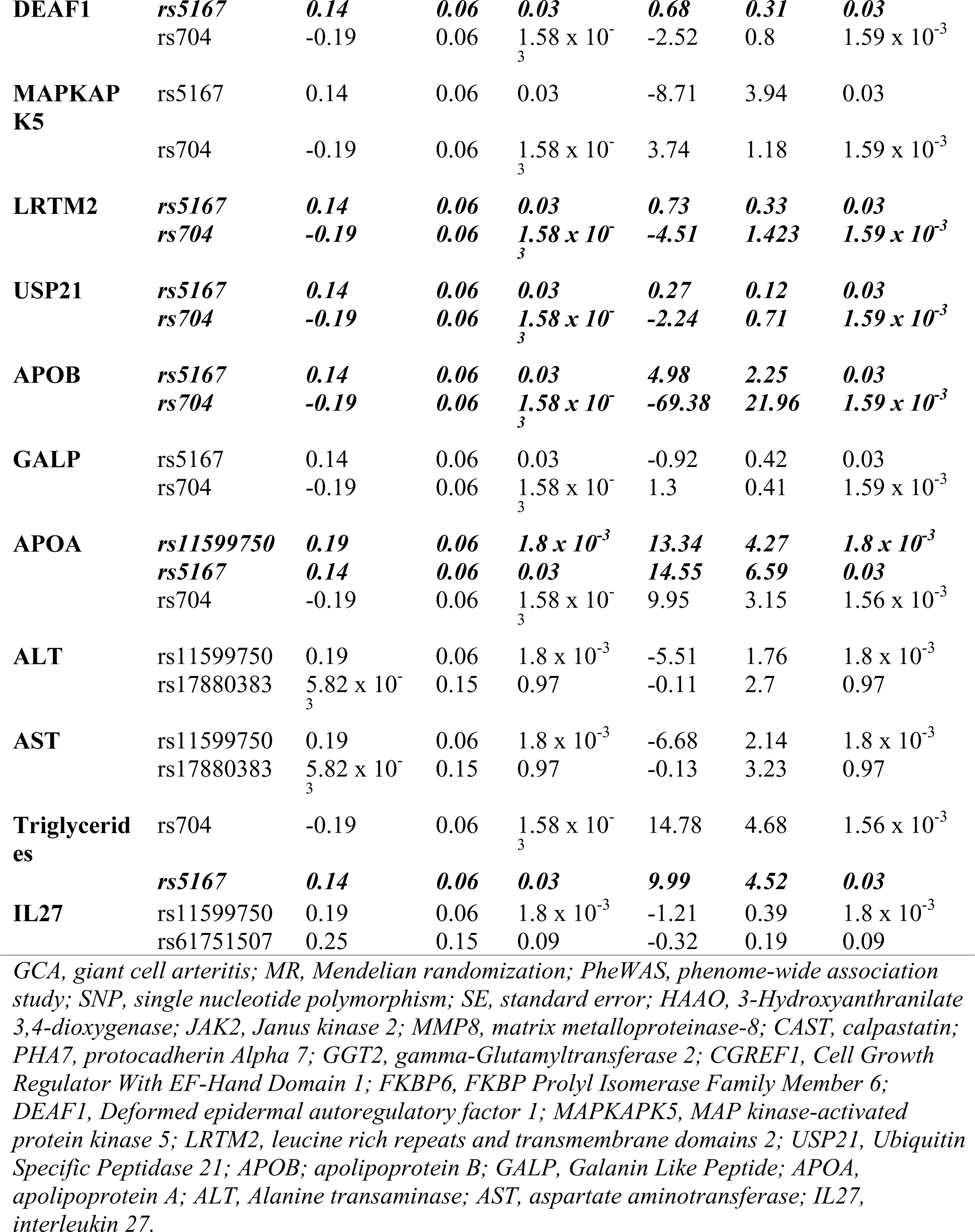
Results of GCA susceptibility MR analyses using proteins/traits identified in PheWAS as exposure variables.

**Supplementary Table 7.** Results of MR-Egger and Cochran’s *Q* sensitivity analyses using the conservative APOL1 PRS with rs704, rs11599750 and rs5167 removed.

| **Risk Score** | **MR-Egger intercept (SE)** | **MR-Egger intercept *P*-value** | **Cochran's *Q*** | **Cochran's *Q* DF** | **Cochran's *Q* *P*-value** |
| --- | --- | --- | --- | --- | --- |
| **Liberal** | -0.01 (0.004) | 0.24 | 2187.22 | 2209 | 0.63 |
| **Intermediate** | 0.03 (0.02) | 0.06 | 80.49 | 101 | 0.93 |
| **Conservative** | -0.13 (0.09) | 0.18 | 5.36 | 7 | 0.62 |

*APOL1*, apolipoprotein-L1; *PRS*, polygenic risk score; *SE*, standard error; *DF*, degrees of freedom.

